# The Amortization of Funding Gene Therapies: Making the “Intangibles” Tangible for Patients

**DOI:** 10.1101/2021.04.16.21255597

**Authors:** Monique Dabbous, Mondher Toumi, Steven Simoens, Juergen Wasem, Yitong Wang, José Huerta Osuna, Clément François, Lieven Annemans, Johann-Matthias Graf von der Schulenburg, Oriol Sola-Morales, Daniel Malone, Louis P. Garrison

## Abstract

Gene replacement therapies (GRTs) are increasingly expected to reach the market. Current healthcare systems are struggling to fund such valuable, costly therapies. GRTs are highly valuable as they extend life through sustained, long-term efficacy or by saving on the costs of a current high-cost standard of care. Some payers have implemented payment models, which do not address the budget impact on the year of acquisition or administration of costly GRTs. This study aims to (1) introduce amortization as an accounting tool within the context of healthcare, specifically for GRTs, (2) present a systematic literature review (SLR) on the amortization or depreciation of pharmaceuticals and medical devices, (3) assess the rationale and feasibility as well as the pros and cons of the amortization of GRTs, and (4) provide recommendations for future steps for the introduction of amortization for GRTs. The limited literature, identified in the SLR, has proposed amortization as a solution for costly, highly valuable GRT funding, but did not fully investigate and detail amortization and its feasibility. This paper further details and illustrates amortization as a promising method for these GRTs by facilitating market and patient access. Current accounting principles and guidelines must evolve to apply amortization to GRTs.

## 1. Introduction

A new class of therapies sometimes referred to as “regenerative medicines” (RMs) has emerged. Defining RMs can be difficult as there is no standard, widely accepted definition. In the United States (US) and the European Union (EU), RMs are generally characterized as therapies dedicated to returning cells, tissues, organs to normal function(1-3). RMs consist of several different types of therapies, including cell therapies, tissue-engineered therapies, and gene therapies (GTs). These therapies are unique in that they depend on the natural regenerative trait of the human body, cells, tissues, and organs. Both the US and the EU have established regulatory pathways to facilitate the market access of RMs(1). One area of RMs, quickly expanding and increasingly gaining attention, is gene replacement therapies (GRTs). GRTs, which directly target the underlying genetic cause of diseases, may replace chronic drugs or even be the first and unique treatment for severe, often rare conditions.

Currently, there are 1,078 clinical trials registered for RMs and advanced therapies, of which 359 are ongoing for GTs as of the first half of 2020 (109 in phase I, 215 in phase II, and 35 in phase III)(4). It is projected that there will be an annual approval rate of 10 new advanced therapy medicinal products by 2030(5). These therapies are among the costliest therapies, are often given in a single administration, and produce large health improvements. Onasemnogene abeparvovec, marketed under the trade name Zolgensma^®^, for the treatment of Spinal Muscular Atrophy (SMA), has been called “the most expensive drug in the world(6)” at $2.1 million for a single administration(7), though some other therapies — notably for hemophilia — often have higher lifetime costs(7).

Despite their high prices, GRTs — in the medium- and long-term — provide high value by extending life through sustained, long-term efficacy or by saving on the costs of a current high-cost standard of care. In anticipation of the arrival of several GTs at high prices for the upfront one-time administration, the issue of what payment model would be necessary to facilitate their market access arises(8). Several countries have already opted to engage in annual installment payment models(8, 9). However, from a budget impact perspective, payers may have to account for the total cost of the therapy in the year of its administration. If payers split the budget impact in line with the installment, they will actually be infringing on the European System Accounting (ESA) Regulation of 2010(10). Therefore, installment payment models only address the payers’ cash flow challenges, but not the challenge of total budget impact, dependent on the time horizon of the budget and budget planning. Such a situation creates resistance to adopting GRTs due to their high budget impact in the first year of administration, which may result in overspending, exceeding the payers’ annual budgets.

As these therapies may provide large long-term benefits, and may replace costly long-term chronic therapies, it would be more appropriate that the budget impact is also spread over the patient’s lifecycle and, hence, that health plans’ budget for more than a year. Current International Financial Reporting Standards (IFRS)(11) and most public accounting regulations and practices treat drugs as “consumable goods.” Therefore, drugs are not eligible for amortization or depreciation over multiple periods. Amortization and depreciation are accounting techniques applied to intangible and tangible assets, respectively, to calculate assets’ values over time. From a formal accounting perspective, spreading the budget impact of GTs over time would be the equivalent to the introduction of amortization for pharmaceuticals, requiring them to be classified as “intangible” assets.

This study aims to (1) introduce the concept of amortization as an accounting tool within the context of healthcare, specifically for GRTs, (2) present a systematic literature review (SLR) on the amortization or depreciation of pharmaceuticals and medical devices, (3) assess the rationale and feasibility as well as the pros and cons of the amortization of GRTs, and (4) provide recommendations for future steps for the introduction of amortization for GRTs.

## 2. Methods

### 2.1 Systematic Literature Review

#### 2.1.1 SLR and Grey Literature Search

A SLR was conducted on February 3^rd^, 2020 via PubMed and grey literature to identify publications and any previously conducted SLRs in the English language relating to the amortization of pharmaceuticals and medical devices. The advanced search strategy is detailed in Table 1. SLR Search Strategy.

A grey literature search was conducted using keywords, including amortization, depreciation, gene therapies, regenerative medicines, pharmaceuticals, medical devices, innovative payment models, tax amortization benefits, tax regulations, mortgage, financing, health economics, health technology agencies, reimbursement, and market access on various news outlets, online search engines, global organizations, governmental agency websites, financial institutions, and economic forums and conferences.

#### 2.2.2 Data Extraction

Publication characteristics, publication details, key discussion points, and full publications were extracted. Additional detail of the SLR publication data extraction table may be referred to in S1. Two independent reviewers analyzed the results in accordance with the search’s predefined inclusion and exclusion criteria. Any discrepancies found between the two reviewers were resolved through discussion and eventually with a third independent reviewer.

### 2.2 Rationale and Feasibility of Amortization

A group of economist experts specialized in pharmaceutical funding elaborated jointly on the rational feasibility as well as pros and cons of amortization via teleconferences and the exchange of documentation and analysis.

## 3. Results

The SLR resulted in 136 publications identified via PubMed. Ultimately, 131 articles were not included in this review. Of these 131 publications excluded, 23 were not available in the English language, their abstracts were available in English, reviewed, and found to meet the exclusion criteria. The remainder of these 131 publications met the exclusion criteria. Thus, only five publications were found to be eligible based on the SLR. The grey literature search yielded two publications. Figure 1 depicts the strategy results in the PRISMA diagram(12, 13).

### 3.1 SLR Publications

The five publications identified in the SLR all mentioned amortization of pharmaceuticals and/or medical devices and were published within the last 12 years. All SLR publication extraction details and data may be referred to in S2.

Blankart, Stargardt, and Schreyögg (2011), mention amortization within the context of reconsidering conditions on granting market exclusivity when the costs of research and development (R&D) are amortized through the use of the active ingredient of the pharmaceutical in treatment of another indication(14). The authors claimed this could result in lower prices for authorization of future indications of the same therapeutic or more specifically of a therapeutic with the same active ingredient(14). In 2008, Sperling, Reulbach, and Kornhuber published a paper briefly mentioning the amortization of investment costs of vagus nerve stimulation therapy equipment. Durham et al. (in 2012) presented a study on the financial benefit of statin therapy or angiotensin therapy for patients with asymptomatic moderate carotid artery stenosis, which included a cost-efficacy model based on amortized costs without detailing the development and implementation of the model and costs(15): it resembled more a discount rate for cost-effectiveness analysis than an amortization scheme. Lastly, in 2014, Cutler et al. published an article titled “Insurance Switching and Mismatch Between the Costs and Benefits of New Technologies(16).” Cutler et al. used economic modelling to study five hypothetical innovative therapies and determined that there existed “disconnects between short-term budget and long-term costs and benefits among initial payers, downstream payers, and society(16).” The authors presented the potential of amortization to be applied to mitigate high upfront costs over time by transferring financial responsibilities among multiple payers over time.

Kleinke and McGee (2015) clearly discussed financing models for innovative therapies, specifically amortization(17). They highlight current challenges with the increasing number of innovative therapies seeking market access in the US, notably the high prices of specialty pharmaceuticals and the challenge of public and private payers seeking to mitigate the impact of their arrival and costs(17). The authors also presented the innovative therapeutic developers’ defense that high prices for such therapies are warranted due to several factors. These included R&D investments, which must be recuperated by large profits and the claim that innovative therapies will prevent future healthcare interventions resulting in cost-savings for payers(17). Lastly, developers argue that these innovative therapies will be the best medical weapon to fight against diseases, therefore, deserving a larger share of the healthcare economy(17). Yet, payers maintain that these prices are simply too high, that some of these products are merely extensions of older products providing marginal benefits, and that, lastly, too many patients are receiving costly therapeutics which do not provide the benefits the patients are seeking.

To reconcile the developers’ and payers’ perspectives, the authors proposed three financial models for consideration including amortization, reinsurance, and rewarding therapy adherence with reduced copayments. All three were said to reduce high upfront costs and to spread them across the healthcare system since in the US, patients change health insurance providers often. The authors defended the implementation of amortization by emphasizing the potential life-long benefits of innovative therapies. If a therapy is meant to extend a patient’s life beyond treatment discontinuation and improve a patient’s quality of life for several years post therapy, then payment and funding schemes of such therapies should reflect such benefits term.

They also suggested that such a model could appeal to US payers due to high insurance provider turnover and the perception that these payers do not benefit from long-term savings. Kleinke and McGee, however, highlighted potential obstacles an amortization payment model may face, including establishing clear definitions, milestones, payment methods, and communication links(17). The article stated that implementation of such a model must be tested with a demonstration project, within a payer-provider system for a specialty product with a well-defined population.

### 3.2 Grey Literature

The grey literature publication extraction details and data may be referred to in S3.

In 2014, Gottlieb and Carino published “Establishing New Payment Provisions for the High Cost of Curing Disease(18).” Gottlieb and Carino proposed that new, innovative payment models involving credit and contracts with amortized costs be established between payers and innovative therapy developers(18). Gottlieb and Carino directly linked amortization to agreements between payers and the industry to facilitate access to these therapies(18).

According to the authors, market access should be rapidly facilitated to encourage developers to engage in the development of therapies. The authors argued that by adopting new and innovative funding frameworks that match the innovative characteristics of such therapies, that their adoption and access will be more efficient. This is because amortization can spread the high upfront cost over time and, in turn, allow for the benefits of the therapy to be recognized(18). Payment terms and a model will have to be developed between the payers and developers so that they may both recognize the benefit of the therapy in the same timeframe. The authors proposed operationalizing amortization via financial agreements with either the developer or another financial intermediary acting as a third party to the transaction by establishing financial terms that allow the payer to record costs in annual increments and for the developer to record revenues in the same manner(18).

The authors highlighted several concerns that may challenge amortization implementation. Payers will be wary as they prefer predictable costs and must optimize their budgets annually. With such new therapies and their long-term benefits, uncertainty may deter payers to participate. Developers will want to enjoy their revenues, return on investment, and high growth rates. Patients suffering from previously incurable diseases will want immediate relief and access to such therapies without having upfront high copayments or out-of-pocket costs as well. Lastly, payer accounting rules will have to adapt in order to implement such new amortization rules(18). The authors describe amortization as a feasible way to address the challenges that these innovative therapies will bring while noting some of the obstacles the accounting technique will have to overcome to be implemented.

A policy brief was published in 2018(19). The authors—Vogler, Paris, and Panteli—presented the policy brief to address policy makers’ concerns with the increasing number of “expensive” pharmaceuticals and the ability to facilitate market access to patients all while fostering and maintaining a sustainable healthcare system(19). The authors mentioned several trends in policy that have been adapted to address innovative therapy issues, including the application of external price referencing (EPR), calls to introduce differential pricing, payers engaging in individual MEAs, and the utilization of other tools, such as health technology assessment (HTA), value-based pricing, horizon scanning, specific funds, tendering, strategic procurement, uptake of biosimilar medicines, cross-collaborations intra-country as well as across countries, and amortization(19). The policy brief recognized that depreciation is already applied in the US healthcare context, but to tangible assets. For example, it is applied to healthcare equipment where payments are made over the time of utilization or the benefit life of the product. The policy brief then questioned whether amortization will actually distract from or delay sustainability and recognized that amortization may help payers manage their budgets, allowing them to fund innovation and facilitate patient access(19).

## 4. Discussion

### 4.1 Amortization

The term depreciation is often misused as interchangeable with amortization. Amortization and depreciation are two similar concepts in accounting, yet, applied to different assets—intangible assets and tangible assets, respectively. Understanding these assets will also be relevant to determining how these concepts and techniques can be applied to healthcare and, specifically, for the financing of GRTs. The following section will elaborate on the inception and definition of amortization as well as briefly make the distinction between the two principles of amortization and depreciation.

#### 4.1.1 Intangible Assets vs. Tangible Assets

Amortization is applied to intangible assets, while depreciation is applied to tangible assets. In accounting, intangible assets are not physical in nature and can range from goodwill, company expertise, and to intellectual property, such as patents, copyrights, and trademarks(20-24).

Intangible assets can represent great value to a company or its owner and they can be the key to a company’s success as well as provide long-term benefits and advantages(22, 25). For example, a patent can be valued at a very high price and can be acquired or transferred between two entities. Intangible assets can be further classified as either “definite” or “indefinite” intangible assets. Additional detail on indefinite vs. definite intangible assets can be referred to in S4.

For accounting purposes, tangible assets are treated similarly to intangible assets, but represent the physical assets a company or entity may possess or acquire(26). They are also noted for their finite life. Tangible assets may include, for example, property, equipment, computers, office supplies, and vehicles. Such assets are recorded on balance sheets at the cost incurred for acquiring them, even though they are the necessary assets that help a company, firm, or business perform efficiently over time.

#### 4.1.2 Amortization and Depreciation

##### 4.1.2-1 Amortization

Amortization is an accounting principle which dates back to the early 1900s. In the US context, Kleinke and McGee trace the use of a mortgage model, as the precursor to modern day amortization, of high costs over time as a relatively simplistic model back to the early 1900s’ US housing market and mortgages. Although the precursor of amortization, this simplistic mortgage model, was applied to a tangible asset (i.e. houses, buildings), modern day amortization has been restricted to intangible assets. In order to stimulate the housing market during the depression, the National Housing Act of 1934 established the Federal Housing Administration and the Federal Savings and Loan Insurance Corporation to ensure mortgages were more accessible to a larger number of Americans by creating and providing more affordable mortgage rates and schemes(27). Ultimately, the Federal Housing Administration was able to succeed as it extended credit via a mortgage model to Americans who otherwise would have been unable to afford to housing(28). Prior to the National Housing Act, mortgages on houses were expected to be paid with a 50% down payment and the rest to be paid over a period of 5 years. With the Act, Mortgages could then be offered with lower down payments and with subsequent payments to be made over a longer period of time, such as 20% down payment and subsequent payments over the next 20-30 years(28). Therefore, Americans were able to become homeowners at a more affordable rate making payments over a longer period and lenders were able to facilitate loans with secured insurance. For additional detail on the history and evolution of amortization refer to S5.

In a more current economic context, amortization is an accounting and financing concept applied to intangible assets. This technique allows for the writing down of a loan or an intangible asset and for repayment to occur via interest and principal payments sufficient to repay the loan in full by its maturity date or on amortization schedule at the end of which the principal in its entirety is due(29, 30). Essentially, the principal amount is the full amount of the loan and the interest is the charged fee for the principal loan(31). Typically, the interest will fall as time progresses while the overall outstanding principal amount left falls as well.

##### 4.1.2-2 Depreciation

The essential difference between amortization and depreciation is that depreciation’s focus is on the nature of deterioration of tangible assets(32). Depreciation is a representation of the decline in value of a tangible asset most likely due to its consumption or obsolescence. In the field of medicine where technology and procedures are ever evolving, certain medical equipment will be rendered outdated when newer versions or models are developed and replace the previous equipment invested in. Indeed, it is the allocation of the cost of a tangible asset over its useful life or life expectancy.

#### 4.1.3 Conducting Amortization and Depreciation

Amortization has been conducted to address high, upfront costs by spreading them over time, over the amortization schedule. This offers the borrower of the loan ample time to pay back the debt or loan afforded for the intangible asset, which allows for the potential value of the intangible asset itself or the potential value it produces to be observed and reaped over time.

The most important reflection of this accounting technique occurs when it can be observed that an asset has shifted from the balance sheet to the income statement of the company. In accounting, there are three primary statements which are highly important in indicating or revealing a company’s performance. These primary statements comprise the balance sheet, the income statement (also known as the profit and loss statement), and the cash flow statement(33, 34). Refer to S6. the Balance Sheet, the Income Statement and the Cash Flow Statement for definitions, details, and calculations.

##### 4.1.3-1 The Application of the Amortization of Intangible Assets: From the Balance Sheet to the Income Statement

Ultimately, recording amortization on the balance sheet and income statement is an accounting technique. On the balance sheet, as a recorded intangible asset is being amortized and used, a decrease in its asset value should be observed(35). Amortization of the recorded intangible asset on the income statement is an expense which reduces net income, as it is a non-cash expense. Therefore, essentially the balance sheet is indicative of the value of the asset being amortized, while the income statement reports on the amortization expenses. The value of the asset should decrease in the balance sheet at different moments in time as time progresses and amortization expenses should also diminish as the time periods they represent also advance (36).

The income statement record of amortization of an intangible asset reflects that the owner of the asset does not have to record it as a large, outstanding balance weighing down the company, potentially constraining their budget and ability to receive credit for their company. This reflects smaller, installment payments, expanding their budgets, and allowing for entities to gain credit elsewhere, have their credit continuously extended, and to conduct business elsewhere in the company(36). As a company or entity pays off the amortization, payments begin to decrease and will reflect positively in the balance sheet and income statement as the amortization expenses diminish.

##### 4.1.3-2 Amortization of Intangible Assets and Tax Deductibles

As companies amortize their intangible assets, payments are typically fixed over the life of an asset and this is reflected on the income statement. An income statement is also used when determining the income tax a company will have to pay per year. A company which is amortizing an intangible asset is often able to claim this annual written off cost as a tax deduction with the amortized amount being the tax deduction reducing the company’s taxable income.

### 4.2 Applicability of Amortization to GRTs

From an accounting perspective, can GRTs qualify as intangible assets? According to the IFRS, intangible assets, other than goodwill, should be individually identifiable, controlled by the company, and deliver future economic benefit. Refer to S7. Intangible Assets vs. Gene Replacement Therapies for details.

It can be argued that innovative medicines are not a physical or capital good, but a rather a proxy for healthcare production as an intangible asset. In this case, amortization could be considered to alleviate budget impact and facilitate market access. Health cannot be purchased and, therefore, healthcare is purchased as its proxy(37). Payers do not buy pharmaceuticals, but rather the information and health production associated with pharmaceutical administration. Essentially, what is purchased is the intangible asset of potential health benefits, it is not a physical asset one can touch, acquire, transfer, or trade. Payers’ rationale in decision-making and purchasing is well-evidenced by the value-based pricing processes established in most countries and driven by health gains associated with the administration of the health technology.

On a balance sheet, numbers recorded should be linked to any asset, intangible or otherwise. The IFRS currently requires accounting standards to expense intangible assets unless there are future economic benefits that the asset produces. Currently, long-term health benefits of GRTs may or may not qualify under IFRS guidelines to be expensed. However, the long-term savings, in terms of less resource utilization related to the disease, avoided loss of productivity, and avoided premium loss to insurance may be considered as future economic benefits produced by the intangible asset. However, the question will arise with whom these benefits will sit, the payer or a third party, and whether these can be reliably measured.

Alternatively, considering the drug as a tangible asset and applying depreciation based on the acquisition price may be explored, even though it does not correspond to the payer’s valuation perspective, it may well be considered as a tangible asset from an accounting perspective. The issue will arise that a tangible asset is a physical asset, but after administration to patients, the asset is no longer physically identifiable. From the developer’s perspective, GRTs may be considered as a tangible physical asset with a well-set price. The question as to whether the same asset can be written on the developer’s balance sheet as a tangible asset and on the insurer’s balance sheet as an intangible asset remains an issue to be clarified.

With respect to the durability of usage, one may well consider that GRTs are a one-time acquisition or administration with a long-term benefit or usage. However, the long-term benefit and long-term usage may not be easily interchangeable from an accounting perspective. In practice, a traditional asset can be traded, but once administered a GRT can no longer be traded. Therefore, if the asset is not capable of being traded, it may have no value from an accounting perspective. The following section sheds light on the complexity of the topic and the challenges when considering whether GRTs could or could not be amortizable. IFRS regulation must evolve and be revised to ensure the eligibility of amortization of GRTs.

#### 4.2.1 The Qualification of GRTs for Amortization in Practice

Assuming GRTs may be considered as intangible assets, certain criteria must be further required to render these eligible for amortization, such as one-time payments and long-term benefits and usage from the perspective of the developing company, allowing for GRTs to be written off as such an asset on the balance sheet. Therefore, the main two challenges for qualifying GRTs for amortization may be reliably quantifying the benefits and determining a time horizon over which these benefits are realized. GRTs are usually a one-time administration product with long- to very long-term benefits. Although evidence and experience may be scarce with approved GRTs, a 10-year period or more durability of efficacy is anticipated so far, and real world-evidence generation are required for as long as 15 years(38). For several GRTs, there are already 7 years of evidence supporting the therapies, which have maintained efficacy. Therefore, considering 10 years may not be unreasonable.

Since GRTs are a heterogeneous class, being a GRT may not be enough to qualify for amortization. Therapies eligible for amortization should be expected to have a short-term administration associated to a delayed dramatic and/or important long-term benefit. Short-term administration, dramatic and important, and long-term benefit must all be clearly defined.

Although one-time administration is clear and straightforward, it might become less clear when considering an oncology product which is administered over one year yet prevents the recurrence of cancer twenty years later. Nevertheless, in the greater perspective of preventing cancer for two decades, the one-year administration may well be considered short-term administration.

The magnitude of the benefit may be obvious with a lifesaving product, while it may be more complicated to establish for a reduction in disability or for an important benefit in a small subgroup of patients. There is a large variability in efficacy between GRTs. It is unclear how long a benefit should be sustained in order to be considered a long-term benefit. The term should be long enough to justify amortization, yet not too long, in order to be credible at time of launch. Not all GRTs may be eligible for amortization and the criteria for efficacy magnitude and duration of effect, should be further developed and defined to identify pharmaceutical products other than GRTs that may qualify for amortization. The definition may encompass products not qualifying as GRTs as the criteria should be product agnostic.

### 4.3 Challenges in the Application of Amortization to GRTs and the Way Forward

Both the SLR and grey literature failed to yield a publication which has a clear definition of amortization from an accounting perspective in the healthcare context. Amortization is often confused with installment payments without considering the budget impact of the costly therapy that should appear in the insurers book on the year of administration. None of the articles detailed specific amortization techniques with implementation obstacles and solutions. Overall, the very limited literature identified endorses amortization as a solution; however, it fails to address the actual payers budget constraint, but rather targets the cash flow, which may not be the main issue for insurers.

From an accounting perspective, an advantage of the application of amortization to innovative health technologies, particularly GRTs, is that payers do not record the full cost of highly priced GRTs on the budget of the year of acquisition on their income statement. Again, amortization has been proposed or mentioned briefly as a potential solution to mitigate high upfront costs(39). However, when it comes to the cash flow, most developers offer installment payment models of usually over a period of 5 years. Current installment payment models do not address the GRT budget impact on the income statement, but rather only the cash flow. If pharmaceuticals are amortized, then the cost of the therapy will be split over several years alleviating the budget impact on the income statement. Refer to Table 2. Advantages of the Amortization of GRTs for GRT Developers and Payers for further benefits of GRT amortization.

It is important to note that even though amortization can aid in controlling the budget impact; in the long-term, insurers may still face sustainability challenges if a large number of these therapies targeting a large population reach the market. Annuity payments are being used already to mitigate high upfront costs and also attempt to assuage the budget impact(40). Jorgensen and Kefalas found that high upfront payments may deter and delay patient access, making it suboptimal for all stakeholders and suggested that annuity-based payments in conjunction with outcomes-based renumeration could be the model which can address both uncertainty and patient access, “without exceeding the net budget impact test(40).” The authors recognized that similar healthcare sustainability challenges which will arise with an increasing number of cell and GTs reaching the market and recognize the need for innovative funding schemes, methods, and strategies to be applied to mitigate the budget impact of these therapies. However, this does not address the high initial budget impact as once used the GRT cost has to be written in full on the balance sheet the year of administration. Therefore, in an annuity-based payment model, the high upfront cost recorded in full on the income statement indicates a large consumption of the budget for that year, being the year of acquisition, and results in a higher budget impact.

Edlin et al. also proposed annuity payments based on leasing principles at cost-effectiveness thresholds, specifically called technology leasing reimbursement scheme(41). This would allow, according to the authors, for costly therapies to gain access while risk would be shared between developers and payers, with upfront payments being mitigated as payments would be spread over time subject to the health technology delivering its claimed health benefit. The authors used trastuzumab (Herceptin^®^) as a case study(41). However, as established before, GRTs cannot be returned, transferred, nor traded once administered, unlike a car or a plane.

The Institute for Clinical and Economic Review (ICER) published a white paper in 2017 based on a Policy Summit meeting in December of 2016(42). The summit focused its discussion onevidence generation, value assessment, and affordability of gene therapies. In addressing affordability, the white paper presented amortization as a “payment strategy” for gene therapies. However, amortization was simply defined as a mechanism mitigating upfront costs by allowing for smaller payments. Furthermore, the summit and white paper discussed amortization as the amortization of loans, with the source of these loans originating from third parties, such as hedge funds, manufacturers, and government. However, all these loans address cash flow issues and do not address budget impact challenges as described above. Similar arguments have been made in publications by Schaffer, Messner, Mestre-Ferrandiz, Tambor, Towse as well as Montazerhodjat, Weinstock, and Lo, where credit mechanisms and healthcare loans, employing amortization, are proposed as solutions to affordability of costly innovative therapies, with various entities assuming financial responsibility, such as government, private investors, and patients as direct consumers(43, 44).

The ICER Membership Policy Summit meeting expressed reservation on the potential impact of these solutions (which are actually loans rather than amortization), claiming that this strategy could result in price increases, delays of underlying issues resulting in future challenges, and the introduction of another party that will impact the cost through its margin.

Reinsurance could be considered as a payment model applicable to costly GRTs, but similar to annuity payments, credit mechanisms, or healthcare loans, it is unable to fully address a high upfront budget impact that falls on the year of administration of the GRT, even when payments are split over years. Reinsurance has been proposed to ensure benefits are shared between payers, especially in fragmented multi-payer systems, such as in the US. It is dedicated to addressing the risk of disproportional distributions of patients treated with such costly therapies and the duration of their engagement with an insurer relative to others. Refer to Table 3.

Payment Models and their ability to address high upfront budget impact for payers, for additional detail on various types of payment models and their implications.

With an increasing number of innovative health technologies reaching the market, the budget impact of GRTs will accumulate over the years and may challenge the sustainability of health insurance or national health services as they will be overwhelmed by “committed budgets related to amortization.” Considering an amortization schedule, this would mean healthcare systems may reach the point at which they can no longer repay the principal amount or cost recorded on the amortization schedule. The aggregate amount of multiple costly GRTs on the market with amortization schemes may become overwhelming. However, it may also contribute to some savings. Amortization may just be the innovative model to employ in such a constrained healthcare system, yet must be contextualized.

It is important to consider two different situations:

1. A GRT replaces an existing chronic costly therapy: in this case, if the price of the GRT is equivalent to the price of the chronic therapies over 5 to 7 years, then beyond 5 to 7 years, insurers will generate savings. Examples of such cases already exist in SMA and hemophilia(45), where lifetime treatment with nusinersen or factor VIII, respectively, is more likely to be avoided. Refer to S8 and S9 for an example in the context of SMA and onasemnogene abeparvovec (Zolgensma^®^).
2. A GRT replaces a cheap therapy or is the first approved therapy: in such an instance, a GRT becomes a new additional expense that may either lead to savings on other healthcare expenditures, such as in Huntington disease, or it creates a new cost with no savings, such as in the case of Dyschromatopsy, where there are currently untreated patients living with disability and there is no treatment. However, this may not be more costly than small molecules or biologics that may be discovered and providing the same level of benefit.

Therefore, it is not obvious at what stage or in what context GRTs will have a net positive or negative budget impact. The evolution and accumulating expertise in the know-how and scientific development of GRTs will likely allow for the reduction in the cost of goods, cost of development, and in the reduction of acquisition budget impact. Finally, budget impact should be put in the perspective of the cost-effectiveness assessment of the GRTs. The time horizon of amortization may be further complicated by the large variability of in the durability of different pharmaceuticals’ efficacy. The interest rate is another variable in facilitating amortization and may easily be adjusted on the discount rate used for national health economics guidelines. The type of amortization model and method may affect the payment schedule. However, this will not be a bottle neck nor obstacle if all previous challenges have been addressed.

Finally, for various amortization methods refer to S10. Amortization Methods and their Application to GRTs. A detailed formula based on the straight-line amortization method is presented in S11. The Straight-Line Amortization Method: Calculating and Modelling. Refer to Table 4. Straight-Line GRT Amortization Table Example, for an example of a straight-line GRT amortization table and its inputs.

Straight-line amortization examples have been conducted using 5 and 10 years of amortization with interest rates of 1% and 3% on approved GTs and CAR-T Cell therapies. The amortization schedules are presented in S12-27. These selected examples clearly show that under various conditions applied, the yearly price of GTs remains clearly below the prices payers are paying for similar drugs that would be a chronic treatment administration for these rare conditions.

However, this may not be true for all GRTs. An example of the model used can be referred to in “Model 1. Model Example: Straight-line GRT Amortization of onasemnogene abeparvovec at a 3% Interest Rate over 10 years.”

#### 4.3.1 Accounting and Tax Regulation Revision

Amortization of GRTs will have an immediate impact on tax reporting of private insurers. Tax legislation is country-specific: hence, country-specific tax legislation adaptation is needed for amortization. In Europe, the EU implements tax-related directives (as opposed to regulations), which still require individual Member States to enact such tax-related directives in their own individual country legislation. The EU does not (at least currently) have a direct role in collecting taxes or setting tax rates. This may well be the case to reimburse the COVID-19 Quantitative Easing Program, a program dedicated to addressing the dramatic impact COVID-19 has had and continues to have major economic structures in the EU in order to “preserve the integrity of the euro area(46).” It is expected that EU will start collecting taxes for reimbursing the quantitative easing program.

Changing accounting and tax regulation country by country at the national level may well be an exhausting process. If one is considering changing accounting at a broader level, it should be done at the Organization for Economic Co-operation and Development (OECD) level. For example, OECD/G20 developed an Inclusive Framework on Base Erosion and Profit Shifting (BEPS), with over 125 countries, which implements 15 actions “to tackle tax avoidance, improve the coherence of international tax rules, and ensure a more transparent tax environment(47).” The OECD has been very effective in driving BEPS. This could be the new avenue to next, or in parallel, address the evolution of IFRS accounting rules to allow amortization of GRTs. The IFRS is developed by the International Accounting Standards Board (IASB), which is the independent, accounting standard-setting body of the IFRS Foundation. The IASB board members meet on a periodic basis and revise the IFRS every 4 years. In the US, Generally Accepted Accounting Principles (GAAP or US-GAAP) are the accounting standard adopted by the US Securities and Exchange Commission, which co-exists with the IFRS. Working concomitantly with US-GAAP and IFRS is probably the best way forward to pursue regulations that allow for the amortization of GRTs.

#### 4.3.2 Eligibility of GRTs for Amortization

The question of defining the eligibility criteria must be addressed at the frontier of clinical pharmacology and health economics. The lack of a clear and unambiguous definition of which GRTs could be eligible for amortization may create a perverse incentive to amortize a broad range of products, while the budget impact spread over several years may be an opportunity for higher prices. A nonprofit organization, such as the Alliance for Regenerative Medicine (ARM), or a scientific society, such as International Society for Pharmacoeconomics and Outcomes Research (ISPOR), could be among several appropriate organizations to set up multidisciplinary task forces to provide recommendations to policy decision makers as to which specifications a medicine should meet to be eligible for amortization.

## 5. Conclusion

In a world where innovative health technology continues to advance, GRTs possess the potential to address unmet needs for patients suffering from rare, severe genetic diseases. As many GRTs are in the pipeline and are expected to be costly, the healthcare system can soon expect the arrival of many GRTs within a short span of time, which payers may struggle to fund them. Currently, healthcare systems across the world are already strained and GRT developers will be pressed to defend the benefits of their therapies, the high valuation of their therapies, and their high price tags assuming robust evidence support that value. Amortization, an accounting principle applied to intangible assets to write off an acquisition cost over time – usually the useful lifetime of the asset – can be applied to GRTs to facilitate market and patient access. Amortization has been mentioned extremely briefly in only a few publications in the literature as a potential innovative method to be applied to GRTs to facilitate market and patient access, until now. However, previous literature has never considered amortization from the feasibility perspective as well as GAAP and IFRS contexts. GRTs can be considered as intangible assets when they are purchased as a proxy of health gain and as payers value them based on the health gains they can produce. Therefore, as intangible assets with medium- to long-term benefits at very high prices, GRTs can be amortized. GRT amortization can allow for payers to avoid high upfront costs and for better budgetary planning.

A seemingly simple straight-line amortization method allows payers and GRT developers the benefit of predictability, payers can predict their budget impact, as straight-line amortization of GRTs results in static payments per payment period and GRT developers will also know their return-on-investment rate. GRT amortization is theoretically and practically feasible, given that national legislation and international guidelines allow for it. GRT amortization still needs to be further investigated to understand what an ideal amortization schedule would be — in terms of the duration and interest rates. Negotiating the duration and interest rates will fall heavily on both parties, GRT developers and payers; however, it is not impossible as complex managed entry agreements and contracting are already occurring between the industry and payers. Trust and clear communication on roles and responsibilities between the two will be critical for the success of GRT amortization. However, third parties such as OECD and IARB, may also play an important role by assessing and facilitating the process.

It is a promising method to open a new door for these innovations by facilitating market and patient access. However, GRT amortization does not come without business-disrupting changes that may happen in parallel and at different levels, which must address the challenges of considering and classifying GRTs as amortizable intangible assets. This will first require international changes at the OECD level and in IFRS accounting. As the US is, by far, the largest market for GRTs and the country with the largest concentration of developers, the evolution and adaptation in accounting rules for GRTs in US-GAAP will be very important. From a broader perspective, it will be critical to define the criteria for pharmaceuticals in general, not only GRTs, to be eligible for amortization, which may be the real challenge that goes beyond GRTs. This must encompass a class of products for chronic conditions that are associated with short-term administration while providing long-term benefits. Such business disruptions will require a joint effort from all stakeholders and a multidisciplinary task force to be engaged to achieve this outcome. Developers, payers, health economists, clinical pharmacologists, and accounting auditors—in close collaboration with financial/accounting regulators at the OECD, IASB and SEC—should be involved to develop and ensure a successful strategy. The amortization of GRTs may well be the most feasible mechanism to enhance a fast access to GRTs at a fair price for all stakeholders and in a sustainable manner for the broader healthcare system. Further investigation in the national and international adoption of the concept will be needed to understand how best to implement GRT amortization.

## Supporting information

Model (Macros disabled to fit MEDRXIV formatting)

## Data Availability

Data reported is easily available via various databases open access to those who wish to replicate the study. Furthermore, all data and methodology are reported on with full transparency.

## Acknowledgements

No funding was involved in the development of this research and manuscript.

## Notes

### Competing Interest Statement

The authors have declared no competing interest.

### Clinical Trial

N/A

### Funding Statement

There was no funding involved in the development of this research and manuscript.

