## Supplementary material for "The Amortization of Funding Gene Therapies: Making the “Intangibles” Tangible for Patients": Table 1

### Table 1. SLR Search Strategy

| **#** | **Topic** | **Searches** | **Results** |
| --- | --- | --- | --- |
| 1 | Amortization, depreciation, reinsurance | (amorti?ation* or amorti?e* or depreciation* or depreciate* or reinsurance*).mp. | 1186 |
| 2 | Cost, model, expenses, annuity | (model* or expense* or rate* or scheme* or payment* or valuation*).mp. | 6284854 |
| 3 |  | cost*.mp. | 682677 |
| 4 |  | (annuity adj2 (payment* or installment*)).mp. | 5 |
| 5 |  | 2 or 3 or 4 | 6728634 |
| 6 |  | 1 and 5 | 747 |
| 7 |  | (pharmaceutical* or drug* or therap*3 or medicine*).mp. | 8953525 |
| 8 |  | (medical adj2 (device* or invention* or instrument*)).mp. | 17757 |
| 9 |  | 6 and 7 | 134 |
| 10 |  | 6 and 8 | 2 |
| **11** | **Final result** | **9 or 10** | **136** |
