## Supplementary material for "The Amortization of Funding Gene Therapies: Making the “Intangibles” Tangible for Patients": Table 2

### Table 2. Advantages of the Amortization of GRTs for GRT Developers and Payers

| Advantages of the Amortization of GRTs for GRT Developers and Payers | |
| --- | --- |
| Developers | Payers |
| - Facilitated market access and patient access - Afforded a timeline to generate the necessary evidence - Ability to experience return on investment and profits over an agreed upon timeline - Reduction in overall economic strain and burden on the healthcare system - Innovative, sustainable model to address payment for innovative therapies | - Facilitated market access and patient access - Mitigate high upfront cost of GRTs - Predictable, agreed upon installment payments over an established timeline - Afforded a timeline to observe evidence addressing - Ability to claim tax deductions based on tax amortization benefits (particularly so for US private payers) - Reduction in overall economic strain and burden on the healthcare system |
