## Supplementary material for "The Amortization of Funding Gene Therapies: Making the “Intangibles” Tangible for Patients": Table 3

### Table 3. Payment Models and their ability to address high upfront budget impact for payers

| Payment Model | Description | Feasibility | Payer Impact | Manufacturer Impact |
| --- | --- | --- | --- | --- |
| Amortization | Amortization is an accounting technique that allows for the writing down of an intangible asset on the balance sheet and for the value to be split over a pre-defined number of successive years according to the amortization schedule This allows to avoid the cost of the asset being concentrated on the year of acquisition. | Amortization can be applied to the cost of Gene Replacement Therapies (GRTs) as intangible assets when generally accepted accounting principles (GAAP) are adapted and evolved.  It will require changes in GAAP and International Financial Reporting Standards (IFRS) to become applicable. | Amortization does for the high upfront budget impact of GRTs to be addressed and mitigated. Amortization allows for the GRT price to be split over several years, rather than be absorbed in the first year of acquisition and administration. | This technique allows for GRT manufacturers to be compensated in full for the therapy at the time of acquisition. |
| Depreciation | Depreciation, similar to amortization, is an accounting technique applied to tangible assets and takes into account the useful life of an asset. The essential difference between amortization and depreciation is that depreciation is applied to tangible assets. When depreciated, a tangible asset should be physically present in the company. | Depreciation is not a feasible payment model strategy as GRTs are not considered as tangible assets by payers. Once a GRT is administered, it is no longer tangible. Depreciation is already applied in the healthcare context when applied to hospitals, healthcare buildings, property, large furniture, and equipment, which are all tangible assets. | N/A | N/A |
| Considering GRT as an “intangible service” | A service is a ready-to-use deliverable that is of value to the customer to perform its activity, such as a background IT infrastructure of an insurer or a website for promoting the insurer image etc. A service is not the core business of the customer, but a tool that may be used to produce the core business. Services could be amortized as intangible asset. | Considering GRTs as a core element of payers’ duties to cover healthcare intervention makes it difficult to consider GRTs as services. | It remains a question for payers to consider if a therapy—more specifically, a GRT—may be considered as a service not core to their business. It does not seem to be the case so far. | It is unlikely that manufacturers will consider a GRT as a service. |
| Leasing Health | Leasing health would mean that a GRT would be paid for by payers for its use, with payments made to the GRT manufacturer on an agreed upon periodic basis. While leasing is common in the healthcare context, for example with large medical equipment (such as MRI machines and surgical robots) which are leased based on their use, its application to GRTs does not fit. Usually, after a lease, medical equipment could be returned or traded in. However, once a GRT is administered, it may not be returned, transferred, or traded in. | Health leasing is not feasible. This is because once administered, a GRT cannot be returned, whereas in commonplace lease agreements, an asset must be returned upon termination of the lease contract. Therefore, even if this is considered as a possible payment model strategy, a payer would not be able to terminate the lease, once initiated. | N/A | N/A |
| Subscription on a yearly basis | If GRTs were subscribed to, a payer would pay fixed costs on a regular basis, for example, annually. Similar to leasing, once administered, GRTs cannot be returned and cannot be discontinued. therefore, the subscription cannot be discontinued. Furthermore, as subscriptions are based on the principle of payment in advance of expected benefits and may be used at different rates by different clients, this cannot be applied to the concept of GRTs. | As with health leasing, subscription would not be feasible for GRTs, for the similar reason that once a subscription ends, services expected are meant to cease as well; however, once administered, GRTs cannot be returned or transferred, and neither can their effects be stopped. While leasing is payment for the use of the GRT, a subscription is an agreement for payment in advance of expected services or outcomes, in both cases neither agreements can be stopped as the GRTs once administered cannot be traded, transferred, returned, reversed, etc. | N/A | N/A |
| Annuity / Installment Payment | Annuity/Installment payments are already being employed in the current healthcare context. Annuity or Installment payments allow for payers to pay for the costs of a GRT on an installment plan, which could be annually or based on another agreed upon schedule between manufacturer and payer. It may be also conditional upon certain criteria, such as outcomes within performance-based agreements. This type of payment model addresses the cash flow but not the budget impact | Annuity or Installment payments are feasible and are already currently implemented for GRTs and other innovative therapies. Therefore, this payment model is feasible and widely used. | Payers are able to address cash flow challenges with this payment model, however, it still does not address the high upfront budget impact of GRTs. Payers are able to make payments on an agreed upon installment schedule and ensure consistent and/or constant appropriate cash flow, but will still record the high upfront budget impact in the year of acquisition and administration of the therapy. | Manufacturers will be able to receive scheduled payments and still be compensated for the therapy in full, according to the payment schedule. However, their revenue is written the year of the sale in their books, while the cash flow differs |
| Reinsurance | Reinsurance occurs when payers insure themselves in the case of large, unpredictable, emergent payouts. Reinsurance payments could occur annually or via an agreed upon timeline. This option rather addresses the risk of disproportional distribution of patients with highly costly therapies among insurers. | If payers consider the risk of disproportional distribution of patients with highly costly therapies among insurers to be a serious issue and GRTs are eligible for reinsurance, this payment model would be feasible. | If GRTs will be eligible to be a part of reinsurance plans, payers will experience a level of protection in unpredictable distributions of the number of patients with highly costly GRTs within their plan. This would be of interest when the number of GRTs and the number of insured patients become very large. Under these circumstances, the risk of disproportional distribution of GRTs is higher. | Manufacturers would not be affected by this payment model. It may facilitate access to patients if payers are less concerned about the risk as they are insured. |
| Healthcoin or Third Party | As a new tradeable currency, healthcoin would convert incremental outcomes produced by a GRT to a common currency. This would appeal to a multi-payer system where there is a high insurance provider turnover. If a payer were to pay for a GRT and the patient were to switch insurance providers, the second or following insurance provider would pay the first for the patient taken on, dependent on healthcoins. | Healthcoin, similar to reinsurance, will be most appealing between payers in a multi-payer system. It could be feasible, once clear definitions and conditions are outlined and abided by all payers engaging in a market utilizing healthcoins. It is unclear if current legal frameworks in several countries would allow for healthcoin to be implemented. This remains to be confirmed | Payers, especially in a multi-payer system, such as in the USA, may find this appealing as they will continue to benefit once a patient, for whom they had provided a GRT, changes health insurance providers. This payment model strategy will not address neither cash flow nor the high upfront budget impact. It does address the disincentive to charge a highly costly therapy where it is perceived that the value may benefit alternative payers. | For the GRT manufacturer, no major direct impact results from the implementation of this payment model. As long as whomever the payer is responsible for the patient receiving the GRTs compensates the therapy, the manufacturer will continue to benefit. It may increase incentives for payers to adopt GRTs |
| Outcomes-Based Agreements | These agreements are already being implemented in healthcare for various types of therapies, including innovative and costly ones. Essentially, these agreements, established between manufacturers and payers, allow for market access of therapies under specific pre-determined, agreed upon conditions directly tied to outcomes these therapies intend to deliver to patients. It may also be associated to installment payment. | This payment model is feasible and already being used with various types of therapies. While these agreements are feasible, there are several major challenges in their implementation, including outcomes follow-up and measuring. Specifically, for therapies, such as GRTs, which provide medium- to long-term benefits, outcomes must be clearly defined prior to engaging in the agreement and their timeline for outcomes follow-up may be much longer than traditional, conventional therapies. | Payers may feel more confident that therapies under outcomes-based agreement schemes they are reimbursing are actually providing the intended benefits to patients. Therefore, it may seem as though payers are not overspending or are not paying for those who do not benefit from the therapy. In reality, this payment model may address cash flow and the effectiveness uncertainty to a certain extent by limiting payment for therapies to specific conditions. However, this payment model does not address the high upfront budget impact as the therapy still is recorded on the books in full in the first year of administration. | Manufacturer impact is relatively low in this payment model. As long as the therapy provides the intended benefits, manufacturers will be compensated for the therapy. In the case where the therapy fails to provide the intended outcomes, manufacturers may, depending on the conditions of the agreement, be required to reimburse payers for lack of intended benefits produced. |
| Consumer Loan | In this payment model, consumers, the patient, are responsible for securing a loan, sometimes referred to as a healthcare loan (HCL), in order to fund their costly therapy. Such a loan could also be amortized, making it more accessible for patients to receive the costly therapy. | Consumer loans are feasible. However, it is not clear whether certain conditions or minimal requirements must be met for a patient to be eligible to secure such a loan. Therefore, there will remain a gap of patients unable to access such a costly therapy. In the case where a patient is able to secure such a loan, payments will cease immediately upon the death of the patient. This means if a patient passes away prematurely, the remainder of the loan will remain unpaid. It is unclear in this scenario who would assume the remaining debt. Furthermore, even if the patient does not receive the intended benefits of the therapy, they would still be committed to repaying their loan. Despite feasibility in theory, this payment model assumes patients pay all or a substantial part of the drug and this will not apply in several countries. It is highly inequitable and loan insurance may be extremely high depending on the patients’ conditions. | This payment model does not impact the payer, payer’s cash flow, or payer’s budget impact as this loan and type of payment model occurs outside the traditional healthcare context. Therefore, it does not contribute to the sustainability of the healthcare system. | Manufacturer impact and risk is high in this payment model. While feasible, if a patient dies, manufacturers will cease to be compensated for the therapy and it is unclear if the remaining loan will be assumed or taken on by another entity. |
| Payer Loan from Private or Government Organization | Similar to consumer loans, payers may also receive loans to fund costly therapies. Payers would be expected to pay back these loans. Payers could receive the loans via various credit mechanism sources, including the government. These loans could also be amortized, as long as the GRTs are considered amortizable. | This payment model could be feasible. It is unclear what would happen in a multi-payer system and whether payment and loan responsibility would switch to a patient’s new provider. If the GAAP and IFRS remain unchanged, drugs will not be amortizable, in which case the drug price will have to be written on the budget of the year of acquisition. This payment model will address cash flow issues. | In the context of a payer loan, cash flow is addressed. However, when it comes to the high upfront budget impact, it will not work as so far drugs are not amortizable. If pharmaceuticals become amortizable, such a loan could be amortized. If this is a loan occurring in a single payer system, the payer may already be the government with their budget already determined by the central government itself. If this type of payment model is taking place in a multi-payer system, such as the USA, it is also unclear how the responsibility of such a loan would shift, once a patient shifts providers. | This payment model will have no impact for manufacturers as they are paid at the time of the therapeutic’s sale. |
| Special Dedicated Governmental Fund | In this payment model case, a government may decide to create special funds to finance GRTs. Such funds exist already, for example, the Cancer Drug Fund, Innovative Medicines Fund, and funding based on diagnosis-related group. Such funds are usually established in single payer systems with their budget established as additional to and separate to the overall health insurance budget. These types of funds are usually financed based on the government’s budget (for example via taxes). It is an artificial way to cover therapeutics within the healthcare system without directly impacting the payer’s budget. | It is feasible and already implemented in single payer systems. It may, however, prove to be more complex to implement in a fragmented payer system, such as in the USA. It may be feasible for the Centers of Medicare & Medicaid Services, if a bill was passed to allow for such a fund to be established. It may be complicated to secure the passing of such a bill as resistance is high to increasing public intervention in healthcare. | For payers, this payment model allows for them to avoid charging the cost of costly therapeutics while still being able to provide access to patients. | Manufacturers in this scheme will still be compensated and they will be paid immediately and in full at the time of therapeutic acquisition by payer. |
| Insurance Pool | This payment model is characterized by several or all health insurers in a catchment area or a country teaming up to contribute to a joint fund in order to finance specific costly projects. In this context, costly GRTs. The contribution depends on the lives covered and may be modulated by some risk factors, such as age, for example. This model addresses the risks of disproportional distribution. | Insurance pooling is feasible and is already implemented, as shown in Germany where payers have created such funds. | Payers mutualize their funds and secure against any negative impact of disproportional distribution of patients requiring GRTs. | Manufacturers may be able to find more payers open to adopting their GRTs as they are covered for disproportional distribution of rare diseases among their policyholders, who require treatment with costly therapies, such as GRTs. |
