## Supplementary material for "The Amortization of Funding Gene Therapies: Making the “Intangibles” Tangible for Patients": Table 4

### Table 4. Straight-Line GRT Amortization Table Example

| Payment Period | Principal Amount Due | Interest Amount Due | Total Payment Amount Due | Principal Balance |
| --- | --- | --- | --- | --- |
| 0 | - | - | - | *Initial Principal Amount will be recorded here. This would be the cost of the GRT* |
| 1 | *The principal amount due for the payment period is calculated by subtracting the interest amount due from the total payment amount due for the payment period (total payment amount due – interest payment amount due).* | *The interest amount due is calculated by multiplying the interest rate per period by the previous balance (r x previous balance). The previous balance for the first payment period (1) is the cost of the GRT.* | *The total payment amount due is the principal amount + the interest amount due.*  *The total payment amount in the straight-line amortization method remains static and uniform throughout all payment periods of the amortization schedule.* | *The new balance or outstanding balance will be calculated by subtracting the principal amount due from the previous balance in the previous payment period. In the case of the first payment period, this would be the cost of the GRT minus the principal amount due in the first payment period (new balance = previous balance – principal amount due).* |
| 2 | *repeated* | *repeated* | *Static, uniform payment* | *repeated* |
| 3 | *repeated* | *repeated* | *Static, uniform payment* | *repeated* |
| … | *repeated* | *repeated* | *Static, uniform payment* | *repeated* |
| … | *repeated* | *repeated* | *Static, uniform payment* | *repeated* |
| … | *repeated* | *repeated* | *Static, uniform payment* | *repeated* |
| 13 | *repeated* | *repeated* | *Static, uniform payment* | *repeated* |
| 14 | *repeated* | *repeated* | *Static, uniform payment* | *repeated* |
| 15 | *repeated* | *repeated* | *Static, uniform payment* | *Ultimately, the calculations are applied throughout the amortization schedule until all payment periods have been paid ending with an outstanding balance of $0, indicating that the principal loan has been paid off in full.* |
