## Supplementary figures and images for "The Amortization of Funding Gene Therapies: Making the “Intangibles” Tangible for Patients"

### Figure

#### Figure 1. SLR Advanced Strategy Results

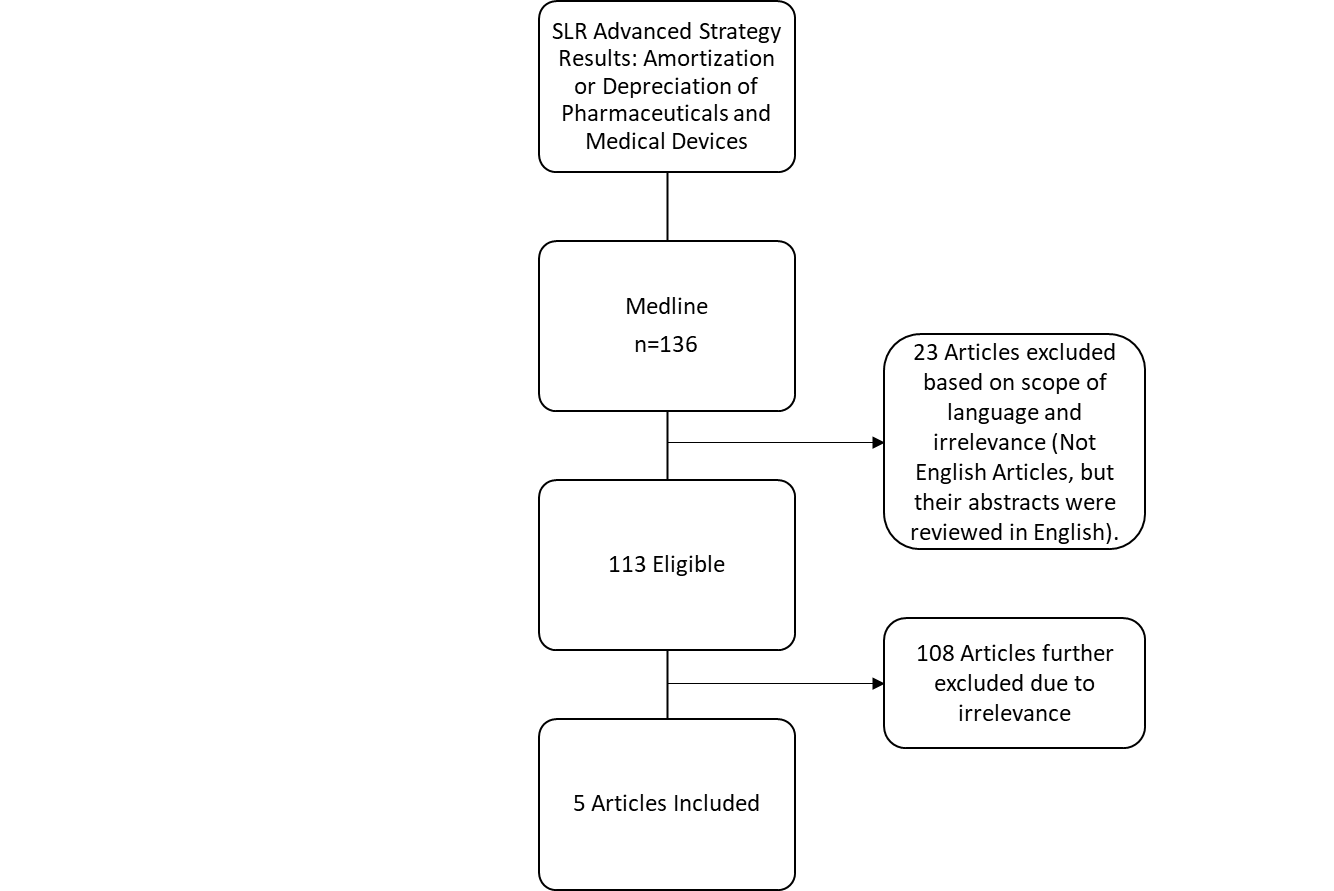
