## Supplementary Materials for "The Amortization of Funding Gene Therapies: Making the “Intangibles” Tangible for Patients"

### Supplementary Material

#### Supplementary 1

Table 1. Example of SLR Data extraction Table

| **Publication Characteristics** | | | | | | | | | | |
| --- | --- | --- | --- | --- | --- | --- | --- | --- | --- | --- |
| **#** | **Year** | **First Author / Institution** | **Title** | **Type of publication** | **Abstract available (y/n)** | **Abstract (copy of abstract)** | **DOI / Link** | **PMID** | **Full text available (y/n)** | **Literature review? (y/n)** |
| 1 |  |  |  |  |  |  |  |  |  |  |

| **Publication details** | | | | | | | | | |  |  |  |  |  |  | **Key Point Discussed in the document (learnings)** |
| --- | --- | --- | --- | --- | --- | --- | --- | --- | --- | --- | --- | --- | --- | --- | --- | --- |
| **to be included?** | | **Region / Country** | | | **Stakeholders** | | | | | **Type of health technology** | | | **Specific disease area ? (Specify)** | **Type of Methodology (specify)** | **Outcomes (y/n) if y, specify type of outcomes** |  |
| **Yes/No** | **Rationale** | **Europe (y/n)** | **US (y/n)** | **Other (specify) (y/n)** | **Patients (y/n)** | **Caregiver/ family (y/n)** | **Society/General population (y/n)** | **HCP (y/n)** | **Decision makers (y/n)** | **Pharmacueticals (y/n)** | **Medical Devices? (y/n)** | **Other? (Specify)** |  |  |  |  |

#### Supplementary 2

Table 2. SLR Publication Details

| **Publication Characteristics** | | | | | | | | | | |
| --- | --- | --- | --- | --- | --- | --- | --- | --- | --- | --- |
| **#** | **Year** | **First Author / Institution** | **Title** | **Type of publication** | **Abstract available (y/n)** | **Abstract (copy of abstract)** | **DOI / Link** | **PMID** | **Full text available (y/n)** | **Literature review? (y/n)** |
| 40 | 2017 | Cutler, David  Ciarametaro, Michael  Long, Genia  Kirson, Noam  Dubois, Robert | Insurance switching and mismatch between the costs and benefits of new technologies. | Journal Article | Y | Abstracts were copy-pasted in this column during the data extraction of the original SLR. Please refer to publication for actual abstract. | N/A | 29261241 | Y | N |
| 47 | 2015 | Kleinke, J D  McGee, Nancy | Breaking the Bank: Three Financing Models for Addressing the Drug Innovation Cost Crisis. [Review] | Journal Article Review | Y | Abstracts were copy-pasted in this column during the data extraction of the original SLR. Please refer to publication for actual abstract. | N/A | 26085900 | Y | N |
| 71 | 2012 | Durham, Christopher Ehlert, Bryan A  Agle, Steven C  Mays, Ashley C  Parker, Frank M  Bogey, William M  Powell, Charles S  Stoner, Michael C | Role of statin therapy and angiotensin blockade in patients with asymptomatic moderate carotid artery stenosis. | Journal Article | Y | Abstracts were copy-pasted in this column during the data extraction of the original SLR. Please refer to publication for actual abstract. | DOI: 10.1016/j.avsg.2011.10.010 | 22285349 | y | N |
| 74 | 2011 | Blankart, Carl Rudolf  Stargardt, Tom  Schreyogg, Jonas | Availability of and access to orphan drugs: an international comparison of pharmaceutical treatments for pulmonary arterial hypertension, Fabry disease, hereditary angioedema and chronic myeloid leukaemia. | Comparative Study  Journal Article  Research Support, Non-U.S. Gov't | y | Abstracts were copy-pasted in this column during the data extraction of the original SLR. Please refer to publication for actual abstract. | doi: 10.2165/11539190-000000000-00000. | 21073206 | y | N |
| 84 | 2009 | Sperling, W  Reulbach, U  Kornhuber, J | Clinical benefits and cost effectiveness of vagus nerve stimulation in a long-term treatment of patients with major depression. | Clinical Trial  Journal Article | y | Abstracts were copy-pasted in this column during the data extraction of the original SLR. Please refer to publication for actual abstract. | N/A | 19452375 | y | n |

| **Publication details** | | | | | | | | | |  |  |  |  |  |  | **Key Point Discussed in the document (learnings)** |
| --- | --- | --- | --- | --- | --- | --- | --- | --- | --- | --- | --- | --- | --- | --- | --- | --- |
| **to be included?** | | **Region / Country** | | | **Stakeholders** | | | | | **Type of health technology** | | | **Specific disease area ? (Specify)** | **Type of Methodology (specify)** | **Outcomes (y/n) if y, specify type of outcomes** |  |
| **Yes/No** | **Rationale** | **Europe (y/n)** | **US (y/n)** | **Other (specify) (y/n)** | **Patients (y/n)** | **Caregiver/ family (y/n)** | **Society/General population (y/n)** | **HCP (y/n)** | **Decision makers (y/n)** | **Pharmacueticals (y/n)** | **Medical Devices? (y/n)** | **Other? (Specify)** |  |  |  |  |
| Y (40) | discusses amortization with models made for gene therapies and 4 others | n | y | N/A | y | n | y | n | y | y | n | N/A | childhood disorder, alzheimer disease, cardiovascular therapy | modelling type | Impact explore was costs of care including on downstream payers in the US context | The article mentions, creative thinking will be require to explore new solutions for payers and developers |
| Y (47) | highly relevant, discusses amortization as a payment model for innovative products | n | y | N/A | N/A | N/A | N/A | N/A | N/A | N/A | N/A | N/A | N/A | N/A | N/A | N/A |
| Y (71) | discusses an amortized cost model for a therapy | n | y | N/A | y | n | y | y | N/A | y | n | N/A | Asymptomatic Moderate Carotid Artery Stenosis | A retrospective review | Amortization details were not mentioned despite amortization costs employed | N/A |
| Y (74) | mention of amortized cost of developing drugs, specifically orphan drugs | y | n | N/A | y | n | y | y | y | y | n | N/A | pulmonary arterial  hypertension (PAH), Fabry disease (FD), hereditary angioedema (HAE) and  chronic myeloid leukaemia (CML) | Comparison | Outcomes here highlighted market authorization variations across countries. | Availability and access play a key role in determining  whether patients will receive adequate  and efficient treatment |
| Y (84) | amortization in context of the electrical impulses delivered allows for amortization of costs | y | n | N/A | y | n | n | y | n | n | y | N/A | major depression | psychopathological ratings and socio-econmic data and comparison | Therefore, the purpose of this study was not only to evaluate the  clinical benefi ts of VNS therapy but also the amortisation of the  initially high investment costs for the equipment (averaging at  present 9 500 S for a VNS system from Cyberonics ™ , Houston,  Texas) within the fi rst 5 years. | In addition to an improvement in  clinical symptoms, the VNS method might en able  an amortisation of costs |

#### Supplementary 3

Grey Literature Publication Details

| **Publication Characteristics** | | | | | | | | | | |
| --- | --- | --- | --- | --- | --- | --- | --- | --- | --- | --- |
| **#** | **Year** | **First Author / Institution** | **Title** | **Type of publication** | **Abstract available (y/n)** | **Abstract (copy of abstract)** | **DOI / Link** | **PMID** | **Full text available (y/n)** | **Literature review? (y/n)** |
| 1a | 2014 | Gottlieb S, Carino T | Establishing new payment provisions for the high cost of curing disease | Article for American Enterprise Institute | y | Abstracts were copy-pasted in this column during the data extraction of the original grey literature search. Please refer to publication for actual abstract. | N/A | N/A | Y | N |
| 2a | 2018 | Vogler S, Paris V, Panteli | Ensuring access to medicines:  How to redesign pricing,  reimbursement and procurement? | Series, a policy brief | y | Abstracts were copy-pasted in this column during the data extraction of the original grey literature search. Please refer to publication for actual abstract. | N/A | N/A | y | N |

| **Publication details** | | | | | | | | | |  |  |  |  |  |  | **Key Point Discussed in the document (learnings)** |
| --- | --- | --- | --- | --- | --- | --- | --- | --- | --- | --- | --- | --- | --- | --- | --- | --- |
| **to be included?** | | **Region / Country** | | | **Stakeholders** | | | | | **Type of health technology** | | | **Specific disease area ? (Specify)** | **Type of Methodology (specify)** | **Outcomes (y/n) if y, specify type of outcomes** |  |
| **Yes/No** | **Rationale** | **Europe (y/n)** | **US (y/n)** | **Other (specify) (y/n)** | **Patients (y/n)** | **Caregiver/ family (y/n)** | **Society/General population (y/n)** | **HCP (y/n)** | **Decision makers (y/n)** | **Pharmacueticals (y/n)** | **Medical Devices? (y/n)** | **Other? (Specify)** |  |  |  |  |
| Y (1a) | discusses amortization as a potential solution to help provide for the cost of these cures for contracting agreements to include costs of treatments to be amortized | n | y | N/A | y | n | y | n | y | y | y | talks about treatments, not necessarily pharmaceuticals or medical devices | N | Discussion | discusses the potential of pricing and payment models for cures | Potential solutions to help provide for the cost of these cures include new credit and contracting arrangements between payers and the health care industry that allow for the cost of treatment to be amortized over many years. |
| Y (2a) | discusses amortization applied to medical equipment but also as a method that can be applied to innovation | y | n | N/A | y | n | y | y | y | y | y | perhaps both but discusses pharmaceuticals, innovative medicines | n | discussion | policy suggestions and recommendations to facilitate payment models for innovative medicines | Policy-makers and purchasers increasingly face very high price tags for new medicines requiring new solutions |

#### Supplementary 4

Indefinite Intangible Assets vs. Definite Intangible Assets

**Indefinite Intangible Assets vs. Definite Intangible Assets**

Indefinite intangible assets can refer to a company’s brand name, for example. A brand name will last with a company indefinitely. Definite intangible assets can refer to potential patents, which expire, or they can refer to time-limited contracts and/or agreements(1). Another interesting characteristic to note about intangible assets in the traditional accounting setting, is that they often work jointly with other assets, specifically tangible assets and, typically, they cannot be valued on a balance sheet independently of other assets. For example, a company’s intangible asset may be its distribution network and the relationships it has built with network stakeholders. However, for a distribution network, this intangible asset brings value with a tangible asset, such as trucks which are able to make deliveries for the distribution network(2,3).

References Supplementary 20

1. Kenton W. Intangible Asset. Online: Investopedia.
2. PENMAN SH. Accounting for Intangible Assets: There is Also an Income Statement. Abacus. 2009; 45: 358-71.
3. Osinski M, Selig P, Matos F, et al. Methods of evaluation of intangible assets and intellectual capital. Journal of Intellectual Capital. 2017; 18: 00-00.

#### Supplementary 5

History and Evolution of Amortization

**History and Evolution of Amortization**

Amortization, in the US, can be traced back to the early 1900s. An article written by Alfred D. Chandler and published in 1913 in The American Economic Review refers to the growing interest in amortization during the early 1900s(1). Chandler references the growing railroad and industrial businesses in the US, which would at the time require from four to five billions of dollars in federal, state, and municipal loans(1). Essentially, the “extinction” or repayment of these loans would be necessary and the best way or method of repaying such large costs would be the subject of Chandler’s article. Chandler references Mr. Hartley Turner of England and his work, which would uphold amortization through sinking funds. Sinking funds are funds formed periodically by an entity to gradually repay a debt or replace a loan taken out. Ultimately, Chandler reviews Turner’s book and addresses how amortization can be used to pay off large loans and proclaims it to be the simplest form of repayment for such large loans, relative to other types of repayment. In 1937, several years after the National Housing Act was passed, the Accounting Review published an article titled “Valuation and Amortization” by Gabriel A.D. Preinreich(2). This article presented definitions of a balance sheet, intangible and tangible assets, and the application of the amortization method. The method of amortization described by Preinreich allows for a true investment not to be directly recorded on a balance sheet, but for the present worth of the asset’s ultimate selling price (total cost) plus its present worth of annuity to be recorded, therefore, spreading the investment or payments over years or a period of time and having the balance sheet reflect this(2). In Preinreich’s article, the useful life of the asset is taken into consideration when calculating and presenting the method of amortization(2).

References Supplementary 21

1. Chandler DA. Amortization. The American Economic Review. 1913; 3: 875-93.
2. Preinreich GAD. Valuation and Amortization. The Accounting Review. 1937; 12: 209-26.

#### Supplementary 6

The Balance Sheet, the Income Statement, and the Cash Flow Statement

**The Balance Sheet:** The balance sheet essentially reports on a company’s total assets, liabilities, and equity. This means that a balance sheet shows what the company has, what it owes, and what is left over at a moment in time – the date stated on the balance sheet. The balance sheet follows an overall simple formula as follows:

Total Assets = Total Liabilities + Equity (1)

The assets on a balance sheet reflect both current and non-current assets(1). Current assets could include cash, trade receivables, investments, and assets held for sale, while non-current assets would include intangible assets, property, equipment, and goodwill(1). Ratios on a balance sheet include, but may not be limited to, debt to equity ratio, inventory turnover, return on net assets, current ratio, and quick ratio(1). These ratios are in the end significant as they allow creditors and lenders to determine whether to continue, extend, or withdraw credit to a company(1).

**The Income Statement:** While the balance sheet illustrates a snapshot of the performance of a company at a moment in time, the income statement reveals how profitable the company actually is over a period of time. The income statement is only indicative of the time period noted on the statement. It is also known as a profit and loss statement and allows for the financial results of a company to be observed, meaning that for the dated period of time, revenues, expenses, gains, and losses are recorded in order to arrive at the final net income of that period, indicating the profitability of the company(2,3) . The main components of an income statement include revenues, expenses, gains and losses, and the net income, but can also usually include, more specifically, tax expenses, post-tax profit or loss for discontinued operations, as well as other comprehensive income(3) . In order to arrive at the net income, the following formula is to be applied:

Net Income = (Revenues + Gains) – (Expenses and Losses)(3)

**The Cash Flow Statement:** The third and final important document of any company or entity is the cash flow statement. Such a statement details the cash flow from firm operations, investing, and financing (4-6). The cash flow statement connects the balance sheet and the income statement (7). Amortization cash flows will appear in the operations’ section of the cash flow statement(8). A cash flow statement is particularly important in assessing the short-term viability of a company or entity as it can be indicative of, for example, the ability of the company to pay the bills(4). Changes in the balance sheet and income statement will affect the cash flow statement and will be reflected on that statement (5). It is considered to be the most important financial statement as a business may look to be performing well on the balance sheet and income statement, yet if it does not have a positive cash flow, the business may not be able to pay its bills, creditors, and employees(5).

References Supplementary 22

1. Tools A. The balance sheet. Online: Accounting Tools, 2018

2. Chen J. Income Statement. Online: Investopedia, 2020.

3. Averkamp H. What is the income statement? Online: Accounting Coach.

4. Murphy C. Understanding the Cash Flow Statement. Online: Investopedia, 2019.

5. Lumen. The Statement of Cash Flows. Online: Lumen - Boundless Accounting.

6. Averkamp H. Cash Flow Statement - Introduction to Cash Flow Statement. Online: Accounting Coach.

7. Barstow SR. Example of How Amortization Affects Financial Statements Online: AZ Central.

8. Ozyasar. What Effect Does Amortization Expense Have on the Statement of Cash Flows? Online: Chron - Small Business.

#### Supplementary 7

Intangible Assets vs. Gene Replacement Therapies

|  | Intangible Assets | Gene Replacement Therapies |
| --- | --- | --- |
| Non-Physical | ✓ | ✓ |
| Represent Great Value | ✓ | ✓ |
| Increase in Value when Used | ✓ | ✓ |
| Provide Long-Term Benefits over a Useful Lifetime | ✓ | ✓ |
| Complex and High Valuation | ✓ | ✓ |
| Unable to be Directly Measured | ✓ | ✓ |

#### Supplementary 8

Cost Comparison and Cost-Savings of Chronic Therapy nusinersen v. Gene Therapy onasemnogene abeparvovec for 1 patient

Cost Comparison and Cost Savings of Chronic Therapy nusinersen v. Gene Therapy onasemnogene abeparvovec for 1 patient

Cost in Dollars

| Year | Cost of Treatment on Chronic Therapy (nusinersen) | Cost-Savings with Gene Therapy |
| --- | --- | --- |
| Y5 | 2250000 | 150000 |
| Y6 | 2625000 | 525000 |
| Y7 | 3000000 | 900000 |
| Y8 | 3375000 | 1275000 |
| Y9 | 3750000 | 1650000 |
| Y10 | 4125000 | 2025000 |
| Y11 | 4500000 | 2400000 |
| Y12 | 4875000 | 2775000 |
| Y13 | 5250000 | 3150000 |
| Y14 | 5625000 | 3525000 |
| Y15 | 6000000 | 3900000 |
| Y16 | 6375000 | 4275000 |
| Y17 | 6750000 | 4650000 |
| Y18 | 7125000 | 5025000 |
| Y19 | 7500000 | 5400000 |
| Y20 | 7875000 | 5775000 |

#### Supplementary 9

Cost-Savings Over 10 Years gene therapy onasemnogene aberparvovec v. chronic therapy nusinersen

|  | onasemnogene abeparvovec | nusinersen |
| --- | --- | --- |
| 1 patient 1 year | 2100000 | 375000 |
| 1 patient 10 years | 2100000 | 4125000 |
| 150 patients 10 years | 315000000 | 618750000 |

*The 1^st^ year of treatment with nusinersen costs $750,000. Every year after that, the yearly cost of nusinersen treatment amounts to $375,000.

#### Supplementary 10

Amortization Methods and their Application to GRTs

**Amortization Methods and their Application to GRTs**

Well-known methods of amortization include the declining balance method, the annuity method, the bullet method, the balloon method, and the increasing balance method. The declining balance method is characterized as an accelerated method, which means that larger payments occur within the earlier years of the amortization schedule and smaller payments are made in the later years until the maturity date is reached and the loan is paid off(1). The annuity method is well-known as it calculates the rate of return of the asset as an investment and also requires the determination of the internal rate of return(2). The bullet method and balloon methods are not usually amortized over the entire amortization schedule. The bullet payment is characterized by only interest payments being made over the amortization schedule with the entire principal amount due at the end of the schedule in a one-time “bullet” payment(3,4). The balloon method is quite similar to the bullet method as there is a large payment involved. However, the balloon method differs as it is characterized by the potential to include smaller installment payments prior to the large “balloon” payment as the remaining large principal balance payment closing off the loan(5). The last type of amortization method is the increasing balance method, also known as negative amortization(6). This method involves unpaid interest and unpaid principal balance being added together, making the overall due balance larger(6). Therefore, in a negative amortization method, the interest payments not made will be added to the loan’s principal balance(6).

When considering which type of amortization method would be most appropriately applied for the amortization of GRTs, several of the aforementioned methods may not be best for the facilitation of market access and patient access of GRTs. For example, the declining balance method could be appealing if it was tied to outcomes. Higher payments could correlate to a higher value a GRT possesses as it may provide larger benefits in the earlier years. However, this would hold true only if the assumption that GRTs administered early enough and produce benefits which are effective in the earlier years post-administration with the benefits beginning to decline over time. Ultimately, the uncertainty around efficacy duration and the complexity of incorporating this into a declining amortization method may prove to be too challenging. The annuity method may also prove too complicated for the beginning of amortizing GRTs as it requires determining and gauging the rate of return of the asset. All remaining three methods, the bullet method, the balloon method, and the increasing balance method, are arguably inappropriate for the amortization of GRTs as they all require significantly high payments by the maturity date of the amortization schedule and, therefore, while they do technically allow for the spreading the acquisition payments of GRTs over time, they would still result in large budget impacts on the healthcare system, and therefore, defeat the purpose of creating an innovative, sustainable payment method.

#### Supplementary 11

The Straight-Line Amortization Method: Calculating and Modelling

**The Straight-Line Amortization Method: Calculating and Modelling**

As it has been established that amortization is conceptually applicable to GRTs as intangible assets, it is necessary to further investigate its practicality and feasibility. There are several methods of amortization. The easiest method of amortization is a straight-line applied amortization method(1,2). The straight-line method of amortization is considered the simplest as it applies the same rate of interest throughout the entire amortization schedule of the intangible asset up until the maturity date of the schedule(1-3). Other well-known methods of amortization include the declining balance method, the annuity method, the bullet method, the balloon method, and the increasing balance method, detailed in Box. 2.

**Straight-Line Amortization Method:** The straight-line method is potentially the best fit method for the amortization of GRTs. This is due to several important reasons. The straight-line method is the simplest and would offer both GRT developers and payers the comfort, stability, and predictability of payments throughout an amortization schedule, liberating a payer’s budget to be redirected towards other resources and investments for public health and allowing for the GRT developers to experience return on investments and profits (2, 4). The uniform installment payments over the GRT amortization schedule allows for the benefits of GRTs to be observed and reaped. This GRT amortization schedule would allow for the GRT developer to further generate critical evidence supporting efficacy duration and safety to address payer uncertainties. Ultimately, the straight-line method offers GRT developers and payers to reconcile their discrepancies to facilitate market and patient access in a sustainable manner for the general healthcare system. Therefore, the straight-line method of amortization was determined to be the preferable method for the application of GRT amortization.

**Calculating and Modelling GRT Amortization:** Several elements must be predetermined prior to applying amortization. One of these major elements is the cost of the GRT, which would represent the principal amount. The remaining critical variables necessary to build and calculate a straight-line amortization model include the amortization schedule which consists of the number of payment periods (payments to be made), and the interest rate per period. With these elements, a GRT amortization schedule detailing payment periods, total payment amount (principal payment + interest payment) per period due, interest payment per period due, principal payment per period due, and overall outstanding balance per period due, can be developed and built for GRT developers and payers to follow.

The simple calculation required for straight-line amortization to determine the payment amount per period of the amortization schedule is the following equation:

$$A=P \frac{r\left( 1+r \right)^{n}}{\left( 1+r \right)^{n}-1}$$

Supplementary 11 Box Continued:

Where: A = Payment Amount per Period, P = Initial Principal (Cost of GRT), r = Interest Rate per Period, and n = Total # of Payments or Periods.

By plugging in the predetermined variables of P, r, and n, the total amount per payment period can be reached, A. The total payment amount due per period, the principal amount + the interest amount due, in the straight-line amortization method remains static and uniform throughout all payment periods of the amortization schedule.

The interest amount due is calculated by multiplying the interest rate per period by the previous balance (r x previous balance). The previous balance for the first payment period is the cost of the GRT, with the remaining previous balances being the balance of the prior payment period.

To determine the principal amount due, the interest amount due is subtracted from the total payment amount due for the payment period (Principal amount due = total payment amount due – interest payment amount due).

To arrive at the new balance amount, the principal amount due is subtracted from the previous balance (new balance or principal balance = previous balance – principal amount due).

#### Supplementary 12

Straight-Line GRT Amortization Table of onasemnogene abeparvovec (Zolgensma®) at an interest rate of 1% monthly over 5 years

| **Payment Period** | **Principal Amount Due** | **Interest Amount Due** | **Total Payment Amount Due** | **Principal Balance** |
| --- | --- | --- | --- | --- |
|  |  |  |  | 2,100,000 |
| 1 | 33,978 | 2,100 | 36,078 | 2,066,022 |
| 2 | 34,012 | 2,066 | 36,078 | 2,032,010 |
| 3 | 34,046 | 2,032 | 36,078 | 1,997,964 |
| 4 | 34,080 | 1,998 | 36,078 | 1,963,884 |
| 5 | 34,114 | 1,964 | 36,078 | 1,929,770 |
| 6 | 34,148 | 1,930 | 36,078 | 1,895,622 |
| 7 | 34,182 | 1,896 | 36,078 | 1,861,439 |
| 8 | 34,217 | 1,861 | 36,078 | 1,827,223 |
| 9 | 34,251 | 1,827 | 36,078 | 1,792,972 |
| 10 | 34,285 | 1,793 | 36,078 | 1,758,687 |
| 11 | 34,319 | 1,759 | 36,078 | 1,724,368 |
| 12 | 34,354 | 1,724 | 36,078 | 1,690,014 |
| 13 | 34,388 | 1,690 | 36,078 | 1,655,626 |
| 14 | 34,422 | 1,656 | 36,078 | 1,621,204 |
| 15 | 34,457 | 1,621 | 36,078 | 1,586,747 |
| 16 | 34,491 | 1,587 | 36,078 | 1,552,256 |
| 17 | 34,526 | 1,552 | 36,078 | 1,517,730 |
| 18 | 34,560 | 1,518 | 36,078 | 1,483,170 |
| 19 | 34,595 | 1,483 | 36,078 | 1,448,575 |
| 20 | 34,629 | 1,449 | 36,078 | 1,413,945 |
| 21 | 34,664 | 1,414 | 36,078 | 1,379,281 |
| 22 | 34,699 | 1,379 | 36,078 | 1,344,583 |
| 23 | 34,733 | 1,345 | 36,078 | 1,309,849 |
| 24 | 34,768 | 1,310 | 36,078 | 1,275,081 |
| 25 | 34,803 | 1,275 | 36,078 | 1,240,278 |
| 26 | 34,838 | 1,240 | 36,078 | 1,205,441 |
| 27 | 34,873 | 1,205 | 36,078 | 1,170,568 |
| 28 | 34,907 | 1,171 | 36,078 | 1,135,661 |
| 29 | 34,942 | 1,136 | 36,078 | 1,100,718 |
| 30 | 34,977 | 1,101 | 36,078 | 1,065,741 |
| 31 | 35,012 | 1,066 | 36,078 | 1,030,729 |
| 32 | 35,047 | 1,031 | 36,078 | 995,681 |
| 33 | 35,082 | 996 | 36,078 | 960,599 |
| 34 | 35,117 | 961 | 36,078 | 925,482 |
| 35 | 35,153 | 925 | 36,078 | 890,329 |
| 36 | 35,188 | 890 | 36,078 | 855,142 |
| 37 | 35,223 | 855 | 36,078 | 819,919 |
| 38 | 35,258 | 820 | 36,078 | 784,661 |
| 39 | 35,293 | 785 | 36,078 | 749,367 |
| 40 | 35,329 | 749 | 36,078 | 714,039 |
| 41 | 35,364 | 714 | 36,078 | 678,675 |
| 42 | 35,399 | 679 | 36,078 | 643,275 |
| 43 | 35,435 | 643 | 36,078 | 607,841 |
| 44 | 35,470 | 608 | 36,078 | 572,371 |
| 45 | 35,506 | 572 | 36,078 | 536,865 |
| 46 | 35,541 | 537 | 36,078 | 501,324 |
| 47 | 35,577 | 501 | 36,078 | 465,747 |
| 48 | 35,612 | 466 | 36,078 | 430,135 |
| 49 | 35,648 | 430 | 36,078 | 394,487 |
| 50 | 35,684 | 394 | 36,078 | 358,804 |
| 51 | 35,719 | 359 | 36,078 | 323,084 |
| 52 | 35,755 | 323 | 36,078 | 287,329 |
| 53 | 35,791 | 287 | 36,078 | 251,539 |
| 54 | 35,826 | 252 | 36,078 | 215,712 |
| 55 | 35,862 | 216 | 36,078 | 179,850 |
| 56 | 35,898 | 180 | 36,078 | 143,952 |
| 57 | 35,934 | 144 | 36,078 | 108,018 |
| 58 | 35,970 | 108 | 36,078 | 72,048 |
| 59 | 36,006 | 72 | 36,078 | 36,042 |
| 60 | 36,042 | 36 | 36,078 | 0 |

#### Supplementary 13

Straight-Line GRT Amortization Table of onasemnogene abeparvovec (Zolgensma®) at an interest rate of 1% monthly over 10 years

| **Payment Period** | **Principal Amount Due** | **Interest Amount Due** | **Total Payment Amount Due** | **Principal Balance** |
| --- | --- | --- | --- | --- |
|  |  |  |  | 2,100,000 |
| 1 | 16,480 | 2,100 | 18,580 | 2,083,520 |
| 2 | 16,496 | 2,084 | 18,580 | 2,067,024 |
| 3 | 16,513 | 2,067 | 18,580 | 2,050,511 |
| 4 | 16,529 | 2,051 | 18,580 | 2,033,982 |
| 5 | 16,546 | 2,034 | 18,580 | 2,017,436 |
| 6 | 16,562 | 2,017 | 18,580 | 2,000,874 |
| 7 | 16,579 | 2,001 | 18,580 | 1,984,295 |
| 8 | 16,595 | 1,984 | 18,580 | 1,967,700 |
| 9 | 16,612 | 1,968 | 18,580 | 1,951,088 |
| 10 | 16,629 | 1,951 | 18,580 | 1,934,459 |
| 11 | 16,645 | 1,934 | 18,580 | 1,917,814 |
| 12 | 16,662 | 1,918 | 18,580 | 1,901,152 |
| 13 | 16,679 | 1,901 | 18,580 | 1,884,473 |
| 14 | 16,695 | 1,884 | 18,580 | 1,867,778 |
| 15 | 16,712 | 1,868 | 18,580 | 1,851,066 |
| 16 | 16,729 | 1,851 | 18,580 | 1,834,337 |
| 17 | 16,745 | 1,834 | 18,580 | 1,817,592 |
| 18 | 16,762 | 1,818 | 18,580 | 1,800,830 |
| 19 | 16,779 | 1,801 | 18,580 | 1,784,051 |
| 20 | 16,796 | 1,784 | 18,580 | 1,767,255 |
| 21 | 16,812 | 1,767 | 18,580 | 1,750,443 |
| 22 | 16,829 | 1,750 | 18,580 | 1,733,614 |
| 23 | 16,846 | 1,734 | 18,580 | 1,716,767 |
| 24 | 16,863 | 1,717 | 18,580 | 1,699,904 |
| 25 | 16,880 | 1,700 | 18,580 | 1,683,025 |
| 26 | 16,897 | 1,683 | 18,580 | 1,666,128 |
| 27 | 16,914 | 1,666 | 18,580 | 1,649,214 |
| 28 | 16,931 | 1,649 | 18,580 | 1,632,284 |
| 29 | 16,947 | 1,632 | 18,580 | 1,615,336 |
| 30 | 16,964 | 1,615 | 18,580 | 1,598,372 |
| 31 | 16,981 | 1,598 | 18,580 | 1,581,391 |
| 32 | 16,998 | 1,581 | 18,580 | 1,564,392 |
| 33 | 17,015 | 1,564 | 18,580 | 1,547,377 |
| 34 | 17,032 | 1,547 | 18,580 | 1,530,345 |
| 35 | 17,049 | 1,530 | 18,580 | 1,513,295 |
| 36 | 17,066 | 1,513 | 18,580 | 1,496,229 |
| 37 | 17,084 | 1,496 | 18,580 | 1,479,145 |
| 38 | 17,101 | 1,479 | 18,580 | 1,462,045 |
| 39 | 17,118 | 1,462 | 18,580 | 1,444,927 |
| 40 | 17,135 | 1,445 | 18,580 | 1,427,792 |
| 41 | 17,152 | 1,428 | 18,580 | 1,410,640 |
| 42 | 17,169 | 1,411 | 18,580 | 1,393,471 |
| 43 | 17,186 | 1,393 | 18,580 | 1,376,285 |
| 44 | 17,203 | 1,376 | 18,580 | 1,359,081 |
| 45 | 17,221 | 1,359 | 18,580 | 1,341,861 |
| 46 | 17,238 | 1,342 | 18,580 | 1,324,623 |
| 47 | 17,255 | 1,325 | 18,580 | 1,307,368 |
| 48 | 17,272 | 1,307 | 18,580 | 1,290,095 |
| 49 | 17,290 | 1,290 | 18,580 | 1,272,806 |
| 50 | 17,307 | 1,273 | 18,580 | 1,255,499 |
| 51 | 17,324 | 1,255 | 18,580 | 1,238,175 |
| 52 | 17,342 | 1,238 | 18,580 | 1,220,833 |
| 53 | 17,359 | 1,221 | 18,580 | 1,203,474 |
| 54 | 17,376 | 1,203 | 18,580 | 1,186,098 |
| 55 | 17,394 | 1,186 | 18,580 | 1,168,704 |
| 56 | 17,411 | 1,169 | 18,580 | 1,151,293 |
| 57 | 17,428 | 1,151 | 18,580 | 1,133,865 |
| 58 | 17,446 | 1,134 | 18,580 | 1,116,419 |
| 59 | 17,463 | 1,116 | 18,580 | 1,098,956 |
| 60 | 17,481 | 1,099 | 18,580 | 1,081,475 |
| 61 | 17,498 | 1,081 | 18,580 | 1,063,977 |
| 62 | 17,516 | 1,064 | 18,580 | 1,046,461 |
| 63 | 17,533 | 1,046 | 18,580 | 1,028,928 |
| 64 | 17,551 | 1,029 | 18,580 | 1,011,377 |
| 65 | 17,568 | 1,011 | 18,580 | 993,808 |
| 66 | 17,586 | 994 | 18,580 | 976,222 |
| 67 | 17,604 | 976 | 18,580 | 958,619 |
| 68 | 17,621 | 959 | 18,580 | 940,998 |
| 69 | 17,639 | 941 | 18,580 | 923,359 |
| 70 | 17,656 | 923 | 18,580 | 905,703 |
| 71 | 17,674 | 906 | 18,580 | 888,029 |
| 72 | 17,692 | 888 | 18,580 | 870,337 |
| 73 | 17,709 | 870 | 18,580 | 852,628 |
| 74 | 17,727 | 853 | 18,580 | 834,900 |
| 75 | 17,745 | 835 | 18,580 | 817,156 |
| 76 | 17,763 | 817 | 18,580 | 799,393 |
| 77 | 17,780 | 799 | 18,580 | 781,613 |
| 78 | 17,798 | 782 | 18,580 | 763,815 |
| 79 | 17,816 | 764 | 18,580 | 745,999 |
| 80 | 17,834 | 746 | 18,580 | 728,165 |
| 81 | 17,852 | 728 | 18,580 | 710,313 |
| 82 | 17,869 | 710 | 18,580 | 692,444 |
| 83 | 17,887 | 692 | 18,580 | 674,557 |
| 84 | 17,905 | 675 | 18,580 | 656,652 |
| 85 | 17,923 | 657 | 18,580 | 638,728 |
| 86 | 17,941 | 639 | 18,580 | 620,787 |
| 87 | 17,959 | 621 | 18,580 | 602,828 |
| 88 | 17,977 | 603 | 18,580 | 584,852 |
| 89 | 17,995 | 585 | 18,580 | 566,857 |
| 90 | 18,013 | 567 | 18,580 | 548,844 |
| 91 | 18,031 | 549 | 18,580 | 530,813 |
| 92 | 18,049 | 531 | 18,580 | 512,764 |
| 93 | 18,067 | 513 | 18,580 | 494,697 |
| 94 | 18,085 | 495 | 18,580 | 476,612 |
| 95 | 18,103 | 477 | 18,580 | 458,509 |
| 96 | 18,121 | 459 | 18,580 | 440,388 |
| 97 | 18,139 | 440 | 18,580 | 422,248 |
| 98 | 18,157 | 422 | 18,580 | 404,091 |
| 99 | 18,176 | 404 | 18,580 | 385,915 |
| 100 | 18,194 | 386 | 18,580 | 367,721 |
| 101 | 18,212 | 368 | 18,580 | 349,509 |
| 102 | 18,230 | 350 | 18,580 | 331,279 |
| 103 | 18,248 | 331 | 18,580 | 313,031 |
| 104 | 18,267 | 313 | 18,580 | 294,764 |
| 105 | 18,285 | 295 | 18,580 | 276,479 |
| 106 | 18,303 | 276 | 18,580 | 258,176 |
| 107 | 18,322 | 258 | 18,580 | 239,854 |
| 108 | 18,340 | 240 | 18,580 | 221,514 |
| 109 | 18,358 | 222 | 18,580 | 203,156 |
| 110 | 18,377 | 203 | 18,580 | 184,780 |
| 111 | 18,395 | 185 | 18,580 | 166,385 |
| 112 | 18,413 | 166 | 18,580 | 147,971 |
| 113 | 18,432 | 148 | 18,580 | 129,539 |
| 114 | 18,450 | 130 | 18,580 | 111,089 |
| 115 | 18,469 | 111 | 18,580 | 92,621 |
| 116 | 18,487 | 93 | 18,580 | 74,134 |
| 117 | 18,506 | 74 | 18,580 | 55,628 |
| 118 | 18,524 | 56 | 18,580 | 37,104 |
| 119 | 18,543 | 37 | 18,580 | 18,561 |
| 120 | 18,561 | 19 | 18,580 | 0 |

#### Supplementary 14

Straight-Line GRT Amortization Table of onasemnogene abeparvovec (Zolgensma®) at an interest rate of 3% monthly over 5 years

| **Payment Period** | **Principal Amount Due** | **Interest Amount Due** | **Total Payment Amount Due** | **Principal Balance** |
| --- | --- | --- | --- | --- |
|  |  |  |  | 2,100,000 |
| 1 | 31,997 | 6,300 | 38,297 | 2,068,003 |
| 2 | 32,093 | 6,204 | 38,297 | 2,035,910 |
| 3 | 32,189 | 6,108 | 38,297 | 2,003,721 |
| 4 | 32,286 | 6,011 | 38,297 | 1,971,436 |
| 5 | 32,382 | 5,914 | 38,297 | 1,939,053 |
| 6 | 32,480 | 5,817 | 38,297 | 1,906,574 |
| 7 | 32,577 | 5,720 | 38,297 | 1,873,997 |
| 8 | 32,675 | 5,622 | 38,297 | 1,841,322 |
| 9 | 32,773 | 5,524 | 38,297 | 1,808,549 |
| 10 | 32,871 | 5,426 | 38,297 | 1,775,678 |
| 11 | 32,970 | 5,327 | 38,297 | 1,742,708 |
| 12 | 33,069 | 5,228 | 38,297 | 1,709,639 |
| 13 | 33,168 | 5,129 | 38,297 | 1,676,472 |
| 14 | 33,267 | 5,029 | 38,297 | 1,643,204 |
| 15 | 33,367 | 4,930 | 38,297 | 1,609,837 |
| 16 | 33,467 | 4,830 | 38,297 | 1,576,370 |
| 17 | 33,568 | 4,729 | 38,297 | 1,542,802 |
| 18 | 33,668 | 4,628 | 38,297 | 1,509,134 |
| 19 | 33,769 | 4,527 | 38,297 | 1,475,364 |
| 20 | 33,871 | 4,426 | 38,297 | 1,441,494 |
| 21 | 33,972 | 4,324 | 38,297 | 1,407,521 |
| 22 | 34,074 | 4,223 | 38,297 | 1,373,447 |
| 23 | 34,176 | 4,120 | 38,297 | 1,339,271 |
| 24 | 34,279 | 4,018 | 38,297 | 1,304,992 |
| 25 | 34,382 | 3,915 | 38,297 | 1,270,610 |
| 26 | 34,485 | 3,812 | 38,297 | 1,236,125 |
| 27 | 34,588 | 3,708 | 38,297 | 1,201,537 |
| 28 | 34,692 | 3,605 | 38,297 | 1,166,844 |
| 29 | 34,796 | 3,501 | 38,297 | 1,132,048 |
| 30 | 34,901 | 3,396 | 38,297 | 1,097,148 |
| 31 | 35,005 | 3,291 | 38,297 | 1,062,142 |
| 32 | 35,110 | 3,186 | 38,297 | 1,027,032 |
| 33 | 35,216 | 3,081 | 38,297 | 991,816 |
| 34 | 35,321 | 2,975 | 38,297 | 956,495 |
| 35 | 35,427 | 2,869 | 38,297 | 921,068 |
| 36 | 35,534 | 2,763 | 38,297 | 885,534 |
| 37 | 35,640 | 2,657 | 38,297 | 849,894 |
| 38 | 35,747 | 2,550 | 38,297 | 814,147 |
| 39 | 35,854 | 2,442 | 38,297 | 778,292 |
| 40 | 35,962 | 2,335 | 38,297 | 742,330 |
| 41 | 36,070 | 2,227 | 38,297 | 706,261 |
| 42 | 36,178 | 2,119 | 38,297 | 670,083 |
| 43 | 36,287 | 2,010 | 38,297 | 633,796 |
| 44 | 36,395 | 1,901 | 38,297 | 597,401 |
| 45 | 36,505 | 1,792 | 38,297 | 560,896 |
| 46 | 36,614 | 1,683 | 38,297 | 524,282 |
| 47 | 36,724 | 1,573 | 38,297 | 487,558 |
| 48 | 36,834 | 1,463 | 38,297 | 450,724 |
| 49 | 36,945 | 1,352 | 38,297 | 413,779 |
| 50 | 37,055 | 1,241 | 38,297 | 376,724 |
| 51 | 37,167 | 1,130 | 38,297 | 339,557 |
| 52 | 37,278 | 1,019 | 38,297 | 302,279 |
| 53 | 37,390 | 907 | 38,297 | 264,889 |
| 54 | 37,502 | 795 | 38,297 | 227,387 |
| 55 | 37,615 | 682 | 38,297 | 189,773 |
| 56 | 37,727 | 569 | 38,297 | 152,045 |
| 57 | 37,841 | 456 | 38,297 | 114,204 |
| 58 | 37,954 | 343 | 38,297 | 76,250 |
| 59 | 38,068 | 229 | 38,297 | 38,182 |
| 60 | 38,182 | 115 | 38,297 | 0 |

#### Supplementary 15

Straight-Line GRT Amortization Table of onasemnogene abeparvovec (Zolgensma®) at an interest rate of 3% monthly over 10 years

| **Payment Period** | **Principal Amount Due** | **Interest Amount Due** | **Total Payment Amount Due** | **Principal Balance** |
| --- | --- | --- | --- | --- |
|  |  |  |  | 2,100,000 |
| 1 | 14,565 | 6,300 | 20,865 | 2,085,435 |
| 2 | 14,608 | 6,256 | 20,865 | 2,070,827 |
| 3 | 14,652 | 6,212 | 20,865 | 2,056,175 |
| 4 | 14,696 | 6,169 | 20,865 | 2,041,479 |
| 5 | 14,740 | 6,124 | 20,865 | 2,026,739 |
| 6 | 14,784 | 6,080 | 20,865 | 2,011,955 |
| 7 | 14,829 | 6,036 | 20,865 | 1,997,126 |
| 8 | 14,873 | 5,991 | 20,865 | 1,982,253 |
| 9 | 14,918 | 5,947 | 20,865 | 1,967,335 |
| 10 | 14,963 | 5,902 | 20,865 | 1,952,372 |
| 11 | 15,007 | 5,857 | 20,865 | 1,937,365 |
| 12 | 15,052 | 5,812 | 20,865 | 1,922,313 |
| 13 | 15,098 | 5,767 | 20,865 | 1,907,215 |
| 14 | 15,143 | 5,722 | 20,865 | 1,892,072 |
| 15 | 15,188 | 5,676 | 20,865 | 1,876,884 |
| 16 | 15,234 | 5,631 | 20,865 | 1,861,650 |
| 17 | 15,280 | 5,585 | 20,865 | 1,846,370 |
| 18 | 15,325 | 5,539 | 20,865 | 1,831,045 |
| 19 | 15,371 | 5,493 | 20,865 | 1,815,673 |
| 20 | 15,418 | 5,447 | 20,865 | 1,800,256 |
| 21 | 15,464 | 5,401 | 20,865 | 1,784,792 |
| 22 | 15,510 | 5,354 | 20,865 | 1,769,282 |
| 23 | 15,557 | 5,308 | 20,865 | 1,753,725 |
| 24 | 15,603 | 5,261 | 20,865 | 1,738,122 |
| 25 | 15,650 | 5,214 | 20,865 | 1,722,472 |
| 26 | 15,697 | 5,167 | 20,865 | 1,706,775 |
| 27 | 15,744 | 5,120 | 20,865 | 1,691,030 |
| 28 | 15,791 | 5,073 | 20,865 | 1,675,239 |
| 29 | 15,839 | 5,026 | 20,865 | 1,659,400 |
| 30 | 15,886 | 4,978 | 20,865 | 1,643,514 |
| 31 | 15,934 | 4,931 | 20,865 | 1,627,580 |
| 32 | 15,982 | 4,883 | 20,865 | 1,611,598 |
| 33 | 16,030 | 4,835 | 20,865 | 1,595,568 |
| 34 | 16,078 | 4,787 | 20,865 | 1,579,490 |
| 35 | 16,126 | 4,738 | 20,865 | 1,563,364 |
| 36 | 16,174 | 4,690 | 20,865 | 1,547,190 |
| 37 | 16,223 | 4,642 | 20,865 | 1,530,967 |
| 38 | 16,272 | 4,593 | 20,865 | 1,514,695 |
| 39 | 16,320 | 4,544 | 20,865 | 1,498,375 |
| 40 | 16,369 | 4,495 | 20,865 | 1,482,005 |
| 41 | 16,419 | 4,446 | 20,865 | 1,465,587 |
| 42 | 16,468 | 4,397 | 20,865 | 1,449,119 |
| 43 | 16,517 | 4,347 | 20,865 | 1,432,602 |
| 44 | 16,567 | 4,298 | 20,865 | 1,416,035 |
| 45 | 16,616 | 4,248 | 20,865 | 1,399,418 |
| 46 | 16,666 | 4,198 | 20,865 | 1,382,752 |
| 47 | 16,716 | 4,148 | 20,865 | 1,366,036 |
| 48 | 16,766 | 4,098 | 20,865 | 1,349,269 |
| 49 | 16,817 | 4,048 | 20,865 | 1,332,453 |
| 50 | 16,867 | 3,997 | 20,865 | 1,315,586 |
| 51 | 16,918 | 3,947 | 20,865 | 1,298,668 |
| 52 | 16,969 | 3,896 | 20,865 | 1,281,699 |
| 53 | 17,019 | 3,845 | 20,865 | 1,264,680 |
| 54 | 17,071 | 3,794 | 20,865 | 1,247,609 |
| 55 | 17,122 | 3,743 | 20,865 | 1,230,488 |
| 56 | 17,173 | 3,691 | 20,865 | 1,213,314 |
| 57 | 17,225 | 3,640 | 20,865 | 1,196,090 |
| 58 | 17,276 | 3,588 | 20,865 | 1,178,814 |
| 59 | 17,328 | 3,536 | 20,865 | 1,161,485 |
| 60 | 17,380 | 3,484 | 20,865 | 1,144,105 |
| 61 | 17,432 | 3,432 | 20,865 | 1,126,673 |
| 62 | 17,485 | 3,380 | 20,865 | 1,109,189 |
| 63 | 17,537 | 3,328 | 20,865 | 1,091,652 |
| 64 | 17,590 | 3,275 | 20,865 | 1,074,062 |
| 65 | 17,642 | 3,222 | 20,865 | 1,056,420 |
| 66 | 17,695 | 3,169 | 20,865 | 1,038,724 |
| 67 | 17,748 | 3,116 | 20,865 | 1,020,976 |
| 68 | 17,802 | 3,063 | 20,865 | 1,003,174 |
| 69 | 17,855 | 3,010 | 20,865 | 985,319 |
| 70 | 17,909 | 2,956 | 20,865 | 967,411 |
| 71 | 17,962 | 2,902 | 20,865 | 949,448 |
| 72 | 18,016 | 2,848 | 20,865 | 931,432 |
| 73 | 18,070 | 2,794 | 20,865 | 913,362 |
| 74 | 18,124 | 2,740 | 20,865 | 895,238 |
| 75 | 18,179 | 2,686 | 20,865 | 877,059 |
| 76 | 18,233 | 2,631 | 20,865 | 858,825 |
| 77 | 18,288 | 2,576 | 20,865 | 840,537 |
| 78 | 18,343 | 2,522 | 20,865 | 822,194 |
| 79 | 18,398 | 2,467 | 20,865 | 803,796 |
| 80 | 18,453 | 2,411 | 20,865 | 785,343 |
| 81 | 18,509 | 2,356 | 20,865 | 766,835 |
| 82 | 18,564 | 2,301 | 20,865 | 748,271 |
| 83 | 18,620 | 2,245 | 20,865 | 729,651 |
| 84 | 18,676 | 2,189 | 20,865 | 710,975 |
| 85 | 18,732 | 2,133 | 20,865 | 692,244 |
| 86 | 18,788 | 2,077 | 20,865 | 673,456 |
| 87 | 18,844 | 2,020 | 20,865 | 654,612 |
| 88 | 18,901 | 1,964 | 20,865 | 635,711 |
| 89 | 18,957 | 1,907 | 20,865 | 616,754 |
| 90 | 19,014 | 1,850 | 20,865 | 597,739 |
| 91 | 19,071 | 1,793 | 20,865 | 578,668 |
| 92 | 19,129 | 1,736 | 20,865 | 559,539 |
| 93 | 19,186 | 1,679 | 20,865 | 540,353 |
| 94 | 19,243 | 1,621 | 20,865 | 521,110 |
| 95 | 19,301 | 1,563 | 20,865 | 501,809 |
| 96 | 19,359 | 1,505 | 20,865 | 482,450 |
| 97 | 19,417 | 1,447 | 20,865 | 463,032 |
| 98 | 19,475 | 1,389 | 20,865 | 443,557 |
| 99 | 19,534 | 1,331 | 20,865 | 424,023 |
| 100 | 19,592 | 1,272 | 20,865 | 404,431 |
| 101 | 19,651 | 1,213 | 20,865 | 384,779 |
| 102 | 19,710 | 1,154 | 20,865 | 365,069 |
| 103 | 19,769 | 1,095 | 20,865 | 345,300 |
| 104 | 19,829 | 1,036 | 20,865 | 325,471 |
| 105 | 19,888 | 976 | 20,865 | 305,583 |
| 106 | 19,948 | 917 | 20,865 | 285,635 |
| 107 | 20,008 | 857 | 20,865 | 265,628 |
| 108 | 20,068 | 797 | 20,865 | 245,560 |
| 109 | 20,128 | 737 | 20,865 | 225,432 |
| 110 | 20,188 | 676 | 20,865 | 205,244 |
| 111 | 20,249 | 616 | 20,865 | 184,995 |
| 112 | 20,310 | 555 | 20,865 | 164,685 |
| 113 | 20,370 | 494 | 20,865 | 144,315 |
| 114 | 20,432 | 433 | 20,865 | 123,883 |
| 115 | 20,493 | 372 | 20,865 | 103,390 |
| 116 | 20,554 | 310 | 20,865 | 82,836 |
| 117 | 20,616 | 249 | 20,865 | 62,220 |
| 118 | 20,678 | 187 | 20,865 | 41,542 |
| 119 | 20,740 | 125 | 20,865 | 20,802 |
| 120 | 20,802 | 62 | 20,865 | 0 |

#### Supplementary 16

Straight-Line GRT Amortization Table of voretigene neparvovec (Luxturna®) at an interest rate of 1% monthly over 5 years

| **Payment Period** | **Principal Amount Due** | **Interest Amount Due** | **Total Payment Amount Due** | **Principal Balance** |
| --- | --- | --- | --- | --- |
|  |  |  |  | 850,000 |
| 1 | 13,753 | 850 | 14,603 | 836,247 |
| 2 | 13,767 | 836 | 14,603 | 822,480 |
| 3 | 13,781 | 822 | 14,603 | 808,700 |
| 4 | 13,794 | 809 | 14,603 | 794,905 |
| 5 | 13,808 | 795 | 14,603 | 781,097 |
| 6 | 13,822 | 781 | 14,603 | 767,275 |
| 7 | 13,836 | 767 | 14,603 | 753,440 |
| 8 | 13,850 | 753 | 14,603 | 739,590 |
| 9 | 13,863 | 740 | 14,603 | 725,727 |
| 10 | 13,877 | 726 | 14,603 | 711,849 |
| 11 | 13,891 | 712 | 14,603 | 697,958 |
| 12 | 13,905 | 698 | 14,603 | 684,053 |
| 13 | 13,919 | 684 | 14,603 | 670,134 |
| 14 | 13,933 | 670 | 14,603 | 656,202 |
| 15 | 13,947 | 656 | 14,603 | 642,255 |
| 16 | 13,961 | 642 | 14,603 | 628,294 |
| 17 | 13,975 | 628 | 14,603 | 614,319 |
| 18 | 13,989 | 614 | 14,603 | 600,331 |
| 19 | 14,003 | 600 | 14,603 | 586,328 |
| 20 | 14,017 | 586 | 14,603 | 572,311 |
| 21 | 14,031 | 572 | 14,603 | 558,281 |
| 22 | 14,045 | 558 | 14,603 | 544,236 |
| 23 | 14,059 | 544 | 14,603 | 530,177 |
| 24 | 14,073 | 530 | 14,603 | 516,104 |
| 25 | 14,087 | 516 | 14,603 | 502,017 |
| 26 | 14,101 | 502 | 14,603 | 487,916 |
| 27 | 14,115 | 488 | 14,603 | 473,801 |
| 28 | 14,129 | 474 | 14,603 | 459,672 |
| 29 | 14,143 | 460 | 14,603 | 445,529 |
| 30 | 14,157 | 446 | 14,603 | 431,371 |
| 31 | 14,172 | 431 | 14,603 | 417,200 |
| 32 | 14,186 | 417 | 14,603 | 403,014 |
| 33 | 14,200 | 403 | 14,603 | 388,814 |
| 34 | 14,214 | 389 | 14,603 | 374,600 |
| 35 | 14,228 | 375 | 14,603 | 360,371 |
| 36 | 14,243 | 360 | 14,603 | 346,129 |
| 37 | 14,257 | 346 | 14,603 | 331,872 |
| 38 | 14,271 | 332 | 14,603 | 317,601 |
| 39 | 14,285 | 318 | 14,603 | 303,315 |
| 40 | 14,300 | 303 | 14,603 | 289,016 |
| 41 | 14,314 | 289 | 14,603 | 274,702 |
| 42 | 14,328 | 275 | 14,603 | 260,373 |
| 43 | 14,343 | 260 | 14,603 | 246,031 |
| 44 | 14,357 | 246 | 14,603 | 231,674 |
| 45 | 14,371 | 232 | 14,603 | 217,302 |
| 46 | 14,386 | 217 | 14,603 | 202,917 |
| 47 | 14,400 | 203 | 14,603 | 188,517 |
| 48 | 14,414 | 189 | 14,603 | 174,102 |
| 49 | 14,429 | 174 | 14,603 | 159,673 |
| 50 | 14,443 | 160 | 14,603 | 145,230 |
| 51 | 14,458 | 145 | 14,603 | 130,772 |
| 52 | 14,472 | 131 | 14,603 | 116,300 |
| 53 | 14,487 | 116 | 14,603 | 101,813 |
| 54 | 14,501 | 102 | 14,603 | 87,312 |
| 55 | 14,516 | 87 | 14,603 | 72,796 |
| 56 | 14,530 | 73 | 14,603 | 58,266 |
| 57 | 14,545 | 58 | 14,603 | 43,722 |
| 58 | 14,559 | 44 | 14,603 | 29,162 |
| 59 | 14,574 | 29 | 14,603 | 14,588 |
| 60 | 14,588 | 15 | 14,603 | 0 |

#### Supplementary 17

Straight-Line GRT Amortization Table of voretigene neparvovec (Luxturna®) at an interest rate of 1% monthly over 10 years

| **Payment Period** | **Principal Amount Due** | **Interest Amount Due** | **Total Payment Amount Due** | **Principal Balance** |
| --- | --- | --- | --- | --- |
|  |  |  |  | 850,000 |
| 1 | 6,670 | 850 | 7,520 | 843,330 |
| 2 | 6,677 | 843 | 7,520 | 836,653 |
| 3 | 6,684 | 837 | 7,520 | 829,969 |
| 4 | 6,690 | 830 | 7,520 | 823,278 |
| 5 | 6,697 | 823 | 7,520 | 816,581 |
| 6 | 6,704 | 817 | 7,520 | 809,878 |
| 7 | 6,710 | 810 | 7,520 | 803,167 |
| 8 | 6,717 | 803 | 7,520 | 796,450 |
| 9 | 6,724 | 796 | 7,520 | 789,726 |
| 10 | 6,731 | 790 | 7,520 | 782,995 |
| 11 | 6,737 | 783 | 7,520 | 776,258 |
| 12 | 6,744 | 776 | 7,520 | 769,514 |
| 13 | 6,751 | 770 | 7,520 | 762,763 |
| 14 | 6,758 | 763 | 7,520 | 756,005 |
| 15 | 6,764 | 756 | 7,520 | 749,241 |
| 16 | 6,771 | 749 | 7,520 | 742,470 |
| 17 | 6,778 | 742 | 7,520 | 735,692 |
| 18 | 6,785 | 736 | 7,520 | 728,907 |
| 19 | 6,791 | 729 | 7,520 | 722,116 |
| 20 | 6,798 | 722 | 7,520 | 715,318 |
| 21 | 6,805 | 715 | 7,520 | 708,513 |
| 22 | 6,812 | 709 | 7,520 | 701,701 |
| 23 | 6,819 | 702 | 7,520 | 694,882 |
| 24 | 6,825 | 695 | 7,520 | 688,057 |
| 25 | 6,832 | 688 | 7,520 | 681,224 |
| 26 | 6,839 | 681 | 7,520 | 674,385 |
| 27 | 6,846 | 674 | 7,520 | 667,539 |
| 28 | 6,853 | 668 | 7,520 | 660,686 |
| 29 | 6,860 | 661 | 7,520 | 653,827 |
| 30 | 6,867 | 654 | 7,520 | 646,960 |
| 31 | 6,873 | 647 | 7,520 | 640,087 |
| 32 | 6,880 | 640 | 7,520 | 633,206 |
| 33 | 6,887 | 633 | 7,520 | 626,319 |
| 34 | 6,894 | 626 | 7,520 | 619,425 |
| 35 | 6,901 | 619 | 7,520 | 612,524 |
| 36 | 6,908 | 613 | 7,520 | 605,616 |
| 37 | 6,915 | 606 | 7,520 | 598,702 |
| 38 | 6,922 | 599 | 7,520 | 591,780 |
| 39 | 6,929 | 592 | 7,520 | 584,851 |
| 40 | 6,936 | 585 | 7,520 | 577,916 |
| 41 | 6,942 | 578 | 7,520 | 570,973 |
| 42 | 6,949 | 571 | 7,520 | 564,024 |
| 43 | 6,956 | 564 | 7,520 | 557,068 |
| 44 | 6,963 | 557 | 7,520 | 550,104 |
| 45 | 6,970 | 550 | 7,520 | 543,134 |
| 46 | 6,977 | 543 | 7,520 | 536,157 |
| 47 | 6,984 | 536 | 7,520 | 529,173 |
| 48 | 6,991 | 529 | 7,520 | 522,181 |
| 49 | 6,998 | 522 | 7,520 | 515,183 |
| 50 | 7,005 | 515 | 7,520 | 508,178 |
| 51 | 7,012 | 508 | 7,520 | 501,166 |
| 52 | 7,019 | 501 | 7,520 | 494,147 |
| 53 | 7,026 | 494 | 7,520 | 487,120 |
| 54 | 7,033 | 487 | 7,520 | 480,087 |
| 55 | 7,040 | 480 | 7,520 | 473,047 |
| 56 | 7,047 | 473 | 7,520 | 466,000 |
| 57 | 7,054 | 466 | 7,520 | 458,945 |
| 58 | 7,061 | 459 | 7,520 | 451,884 |
| 59 | 7,068 | 452 | 7,520 | 444,815 |
| 60 | 7,076 | 445 | 7,520 | 437,740 |
| 61 | 7,083 | 438 | 7,520 | 430,657 |
| 62 | 7,090 | 431 | 7,520 | 423,567 |
| 63 | 7,097 | 424 | 7,520 | 416,471 |
| 64 | 7,104 | 416 | 7,520 | 409,367 |
| 65 | 7,111 | 409 | 7,520 | 402,256 |
| 66 | 7,118 | 402 | 7,520 | 395,138 |
| 67 | 7,125 | 395 | 7,520 | 388,012 |
| 68 | 7,132 | 388 | 7,520 | 380,880 |
| 69 | 7,139 | 381 | 7,520 | 373,741 |
| 70 | 7,147 | 374 | 7,520 | 366,594 |
| 71 | 7,154 | 367 | 7,520 | 359,440 |
| 72 | 7,161 | 359 | 7,520 | 352,279 |
| 73 | 7,168 | 352 | 7,520 | 345,111 |
| 74 | 7,175 | 345 | 7,520 | 337,936 |
| 75 | 7,182 | 338 | 7,520 | 330,753 |
| 76 | 7,190 | 331 | 7,520 | 323,564 |
| 77 | 7,197 | 324 | 7,520 | 316,367 |
| 78 | 7,204 | 316 | 7,520 | 309,163 |
| 79 | 7,211 | 309 | 7,520 | 301,952 |
| 80 | 7,218 | 302 | 7,520 | 294,733 |
| 81 | 7,226 | 295 | 7,520 | 287,508 |
| 82 | 7,233 | 288 | 7,520 | 280,275 |
| 83 | 7,240 | 280 | 7,520 | 273,035 |
| 84 | 7,247 | 273 | 7,520 | 265,788 |
| 85 | 7,255 | 266 | 7,520 | 258,533 |
| 86 | 7,262 | 259 | 7,520 | 251,271 |
| 87 | 7,269 | 251 | 7,520 | 244,002 |
| 88 | 7,276 | 244 | 7,520 | 236,726 |
| 89 | 7,284 | 237 | 7,520 | 229,442 |
| 90 | 7,291 | 229 | 7,520 | 222,151 |
| 91 | 7,298 | 222 | 7,520 | 214,853 |
| 92 | 7,306 | 215 | 7,520 | 207,547 |
| 93 | 7,313 | 208 | 7,520 | 200,235 |
| 94 | 7,320 | 200 | 7,520 | 192,914 |
| 95 | 7,327 | 193 | 7,520 | 185,587 |
| 96 | 7,335 | 186 | 7,520 | 178,252 |
| 97 | 7,342 | 178 | 7,520 | 170,910 |
| 98 | 7,349 | 171 | 7,520 | 163,561 |
| 99 | 7,357 | 164 | 7,520 | 156,204 |
| 100 | 7,364 | 156 | 7,520 | 148,840 |
| 101 | 7,372 | 149 | 7,520 | 141,468 |
| 102 | 7,379 | 141 | 7,520 | 134,089 |
| 103 | 7,386 | 134 | 7,520 | 126,703 |
| 104 | 7,394 | 127 | 7,520 | 119,309 |
| 105 | 7,401 | 119 | 7,520 | 111,908 |
| 106 | 7,408 | 112 | 7,520 | 104,500 |
| 107 | 7,416 | 104 | 7,520 | 97,084 |
| 108 | 7,423 | 97 | 7,520 | 89,661 |
| 109 | 7,431 | 90 | 7,520 | 82,230 |
| 110 | 7,438 | 82 | 7,520 | 74,792 |
| 111 | 7,446 | 75 | 7,520 | 67,346 |
| 112 | 7,453 | 67 | 7,520 | 59,893 |
| 113 | 7,460 | 60 | 7,520 | 52,433 |
| 114 | 7,468 | 52 | 7,520 | 44,965 |
| 115 | 7,475 | 45 | 7,520 | 37,489 |
| 116 | 7,483 | 37 | 7,520 | 30,006 |
| 117 | 7,490 | 30 | 7,520 | 22,516 |
| 118 | 7,498 | 23 | 7,520 | 15,018 |
| 119 | 7,505 | 15 | 7,520 | 7,513 |
| 120 | 7,513 | 8 | 7,520 | 0 |

#### Supplementary 18

Straight-Line GRT Amortization Table of voretigene neparvovec (Luxturna®) at an interest rate of 3% monthly over 5 years

| **Payment Period** | **Principal Amount Due** | **Interest Amount Due** | **Total Payment Amount Due** | **Principal Balance** |
| --- | --- | --- | --- | --- |
|  |  |  |  | 850,000 |
| 1 | 12,951 | 2,550 | 15,501 | 837,049 |
| 2 | 12,990 | 2,511 | 15,501 | 824,059 |
| 3 | 13,029 | 2,472 | 15,501 | 811,030 |
| 4 | 13,068 | 2,433 | 15,501 | 797,962 |
| 5 | 13,107 | 2,394 | 15,501 | 784,855 |
| 6 | 13,147 | 2,355 | 15,501 | 771,708 |
| 7 | 13,186 | 2,315 | 15,501 | 758,522 |
| 8 | 13,226 | 2,276 | 15,501 | 745,297 |
| 9 | 13,265 | 2,236 | 15,501 | 732,032 |
| 10 | 13,305 | 2,196 | 15,501 | 718,727 |
| 11 | 13,345 | 2,156 | 15,501 | 705,382 |
| 12 | 13,385 | 2,116 | 15,501 | 691,997 |
| 13 | 13,425 | 2,076 | 15,501 | 678,572 |
| 14 | 13,465 | 2,036 | 15,501 | 665,106 |
| 15 | 13,506 | 1,995 | 15,501 | 651,601 |
| 16 | 13,546 | 1,955 | 15,501 | 638,054 |
| 17 | 13,587 | 1,914 | 15,501 | 624,468 |
| 18 | 13,628 | 1,873 | 15,501 | 610,840 |
| 19 | 13,669 | 1,833 | 15,501 | 597,171 |
| 20 | 13,710 | 1,792 | 15,501 | 583,462 |
| 21 | 13,751 | 1,750 | 15,501 | 569,711 |
| 22 | 13,792 | 1,709 | 15,501 | 555,919 |
| 23 | 13,833 | 1,668 | 15,501 | 542,086 |
| 24 | 13,875 | 1,626 | 15,501 | 528,211 |
| 25 | 13,916 | 1,585 | 15,501 | 514,295 |
| 26 | 13,958 | 1,543 | 15,501 | 500,336 |
| 27 | 14,000 | 1,501 | 15,501 | 486,336 |
| 28 | 14,042 | 1,459 | 15,501 | 472,294 |
| 29 | 14,084 | 1,417 | 15,501 | 458,210 |
| 30 | 14,126 | 1,375 | 15,501 | 444,084 |
| 31 | 14,169 | 1,332 | 15,501 | 429,915 |
| 32 | 14,211 | 1,290 | 15,501 | 415,703 |
| 33 | 14,254 | 1,247 | 15,501 | 401,449 |
| 34 | 14,297 | 1,204 | 15,501 | 387,153 |
| 35 | 14,340 | 1,161 | 15,501 | 372,813 |
| 36 | 14,383 | 1,118 | 15,501 | 358,430 |
| 37 | 14,426 | 1,075 | 15,501 | 344,005 |
| 38 | 14,469 | 1,032 | 15,501 | 329,536 |
| 39 | 14,512 | 989 | 15,501 | 315,023 |
| 40 | 14,556 | 945 | 15,501 | 300,467 |
| 41 | 14,600 | 901 | 15,501 | 285,867 |
| 42 | 14,643 | 858 | 15,501 | 271,224 |
| 43 | 14,687 | 814 | 15,501 | 256,537 |
| 44 | 14,731 | 770 | 15,501 | 241,805 |
| 45 | 14,776 | 725 | 15,501 | 227,029 |
| 46 | 14,820 | 681 | 15,501 | 212,209 |
| 47 | 14,864 | 637 | 15,501 | 197,345 |
| 48 | 14,909 | 592 | 15,501 | 182,436 |
| 49 | 14,954 | 547 | 15,501 | 167,482 |
| 50 | 14,999 | 502 | 15,501 | 152,483 |
| 51 | 15,044 | 457 | 15,501 | 137,440 |
| 52 | 15,089 | 412 | 15,501 | 122,351 |
| 53 | 15,134 | 367 | 15,501 | 107,217 |
| 54 | 15,179 | 322 | 15,501 | 92,038 |
| 55 | 15,225 | 276 | 15,501 | 76,813 |
| 56 | 15,271 | 230 | 15,501 | 61,542 |
| 57 | 15,316 | 185 | 15,501 | 46,226 |
| 58 | 15,362 | 139 | 15,501 | 30,863 |
| 59 | 15,408 | 93 | 15,501 | 15,455 |
| 60 | 15,455 | 46 | 15,501 | 0 |

#### Supplementary 19

Straight-Line GRT Amortization Table of voretigene neparvovec (Luxturna®) at an interest rate of 3% monthly over 10 years

| **Payment Period** | **Principal Amount Due** | **Interest Amount Due** | **Total Payment Amount Due** | **Principal Balance** |
| --- | --- | --- | --- | --- |
|  |  |  |  | 850,000 |
| 1 | 5,895 | 2,550 | 8,445 | 844,105 |
| 2 | 5,913 | 2,532 | 8,445 | 838,192 |
| 3 | 5,931 | 2,515 | 8,445 | 832,261 |
| 4 | 5,948 | 2,497 | 8,445 | 826,313 |
| 5 | 5,966 | 2,479 | 8,445 | 820,347 |
| 6 | 5,984 | 2,461 | 8,445 | 814,363 |
| 7 | 6,002 | 2,443 | 8,445 | 808,361 |
| 8 | 6,020 | 2,425 | 8,445 | 802,340 |
| 9 | 6,038 | 2,407 | 8,445 | 796,302 |
| 10 | 6,056 | 2,389 | 8,445 | 790,246 |
| 11 | 6,074 | 2,371 | 8,445 | 784,172 |
| 12 | 6,093 | 2,353 | 8,445 | 778,079 |
| 13 | 6,111 | 2,334 | 8,445 | 771,968 |
| 14 | 6,129 | 2,316 | 8,445 | 765,839 |
| 15 | 6,148 | 2,298 | 8,445 | 759,691 |
| 16 | 6,166 | 2,279 | 8,445 | 753,525 |
| 17 | 6,185 | 2,261 | 8,445 | 747,340 |
| 18 | 6,203 | 2,242 | 8,445 | 741,137 |
| 19 | 6,222 | 2,223 | 8,445 | 734,915 |
| 20 | 6,240 | 2,205 | 8,445 | 728,675 |
| 21 | 6,259 | 2,186 | 8,445 | 722,416 |
| 22 | 6,278 | 2,167 | 8,445 | 716,138 |
| 23 | 6,297 | 2,148 | 8,445 | 709,841 |
| 24 | 6,316 | 2,130 | 8,445 | 703,526 |
| 25 | 6,335 | 2,111 | 8,445 | 697,191 |
| 26 | 6,354 | 2,092 | 8,445 | 690,837 |
| 27 | 6,373 | 2,073 | 8,445 | 684,465 |
| 28 | 6,392 | 2,053 | 8,445 | 678,073 |
| 29 | 6,411 | 2,034 | 8,445 | 671,662 |
| 30 | 6,430 | 2,015 | 8,445 | 665,232 |
| 31 | 6,449 | 1,996 | 8,445 | 658,782 |
| 32 | 6,469 | 1,976 | 8,445 | 652,313 |
| 33 | 6,488 | 1,957 | 8,445 | 645,825 |
| 34 | 6,508 | 1,937 | 8,445 | 639,317 |
| 35 | 6,527 | 1,918 | 8,445 | 632,790 |
| 36 | 6,547 | 1,898 | 8,445 | 626,243 |
| 37 | 6,566 | 1,879 | 8,445 | 619,677 |
| 38 | 6,586 | 1,859 | 8,445 | 613,091 |
| 39 | 6,606 | 1,839 | 8,445 | 606,485 |
| 40 | 6,626 | 1,819 | 8,445 | 599,859 |
| 41 | 6,646 | 1,800 | 8,445 | 593,214 |
| 42 | 6,666 | 1,780 | 8,445 | 586,548 |
| 43 | 6,686 | 1,760 | 8,445 | 579,863 |
| 44 | 6,706 | 1,740 | 8,445 | 573,157 |
| 45 | 6,726 | 1,719 | 8,445 | 566,431 |
| 46 | 6,746 | 1,699 | 8,445 | 559,685 |
| 47 | 6,766 | 1,679 | 8,445 | 552,919 |
| 48 | 6,786 | 1,659 | 8,445 | 546,133 |
| 49 | 6,807 | 1,638 | 8,445 | 539,326 |
| 50 | 6,827 | 1,618 | 8,445 | 532,499 |
| 51 | 6,848 | 1,597 | 8,445 | 525,651 |
| 52 | 6,868 | 1,577 | 8,445 | 518,783 |
| 53 | 6,889 | 1,556 | 8,445 | 511,894 |
| 54 | 6,909 | 1,536 | 8,445 | 504,985 |
| 55 | 6,930 | 1,515 | 8,445 | 498,054 |
| 56 | 6,951 | 1,494 | 8,445 | 491,103 |
| 57 | 6,972 | 1,473 | 8,445 | 484,132 |
| 58 | 6,993 | 1,452 | 8,445 | 477,139 |
| 59 | 7,014 | 1,431 | 8,445 | 470,125 |
| 60 | 7,035 | 1,410 | 8,445 | 463,090 |
| 61 | 7,056 | 1,389 | 8,445 | 456,034 |
| 62 | 7,077 | 1,368 | 8,445 | 448,957 |
| 63 | 7,098 | 1,347 | 8,445 | 441,859 |
| 64 | 7,120 | 1,326 | 8,445 | 434,739 |
| 65 | 7,141 | 1,304 | 8,445 | 427,598 |
| 66 | 7,162 | 1,283 | 8,445 | 420,436 |
| 67 | 7,184 | 1,261 | 8,445 | 413,252 |
| 68 | 7,205 | 1,240 | 8,445 | 406,047 |
| 69 | 7,227 | 1,218 | 8,445 | 398,820 |
| 70 | 7,249 | 1,196 | 8,445 | 391,571 |
| 71 | 7,270 | 1,175 | 8,445 | 384,301 |
| 72 | 7,292 | 1,153 | 8,445 | 377,008 |
| 73 | 7,314 | 1,131 | 8,445 | 369,694 |
| 74 | 7,336 | 1,109 | 8,445 | 362,358 |
| 75 | 7,358 | 1,087 | 8,445 | 355,000 |
| 76 | 7,380 | 1,065 | 8,445 | 347,620 |
| 77 | 7,402 | 1,043 | 8,445 | 340,217 |
| 78 | 7,425 | 1,021 | 8,445 | 332,793 |
| 79 | 7,447 | 998 | 8,445 | 325,346 |
| 80 | 7,469 | 976 | 8,445 | 317,877 |
| 81 | 7,492 | 954 | 8,445 | 310,385 |
| 82 | 7,514 | 931 | 8,445 | 302,871 |
| 83 | 7,537 | 909 | 8,445 | 295,335 |
| 84 | 7,559 | 886 | 8,445 | 287,776 |
| 85 | 7,582 | 863 | 8,445 | 280,194 |
| 86 | 7,605 | 841 | 8,445 | 272,589 |
| 87 | 7,627 | 818 | 8,445 | 264,962 |
| 88 | 7,650 | 795 | 8,445 | 257,312 |
| 89 | 7,673 | 772 | 8,445 | 249,638 |
| 90 | 7,696 | 749 | 8,445 | 241,942 |
| 91 | 7,719 | 726 | 8,445 | 234,223 |
| 92 | 7,743 | 703 | 8,445 | 226,480 |
| 93 | 7,766 | 679 | 8,445 | 218,714 |
| 94 | 7,789 | 656 | 8,445 | 210,925 |
| 95 | 7,812 | 633 | 8,445 | 203,113 |
| 96 | 7,836 | 609 | 8,445 | 195,277 |
| 97 | 7,859 | 586 | 8,445 | 187,418 |
| 98 | 7,883 | 562 | 8,445 | 179,535 |
| 99 | 7,907 | 539 | 8,445 | 171,628 |
| 100 | 7,930 | 515 | 8,445 | 163,698 |
| 101 | 7,954 | 491 | 8,445 | 155,744 |
| 102 | 7,978 | 467 | 8,445 | 147,766 |
| 103 | 8,002 | 443 | 8,445 | 139,764 |
| 104 | 8,026 | 419 | 8,445 | 131,738 |
| 105 | 8,050 | 395 | 8,445 | 123,688 |
| 106 | 8,074 | 371 | 8,445 | 115,614 |
| 107 | 8,098 | 347 | 8,445 | 107,516 |
| 108 | 8,123 | 323 | 8,445 | 99,393 |
| 109 | 8,147 | 298 | 8,445 | 91,246 |
| 110 | 8,171 | 274 | 8,445 | 83,075 |
| 111 | 8,196 | 249 | 8,445 | 74,879 |
| 112 | 8,221 | 225 | 8,445 | 66,658 |
| 113 | 8,245 | 200 | 8,445 | 58,413 |
| 114 | 8,270 | 175 | 8,445 | 50,143 |
| 115 | 8,295 | 150 | 8,445 | 41,848 |
| 116 | 8,320 | 126 | 8,445 | 33,529 |
| 117 | 8,345 | 101 | 8,445 | 25,184 |
| 118 | 8,370 | 76 | 8,445 | 16,815 |
| 119 | 8,395 | 50 | 8,445 | 8,420 |
| 120 | 8,420 | 25 | 8,445 | 0 |

#### Supplementary 20

Straight-Line GRT Amortization Table of axicabtagene ciloleucel (Yescarta®) at an interest rate of 1% monthly over 5 years

| **Payment Period** | **Principal Amount Due** | **Interest Amount Due** | **Total Payment Amount Due** | **Principal Balance** |
| --- | --- | --- | --- | --- |
|  |  |  |  | 373,000 |
| 1 | 6,035 | 373 | 6,408 | 366,965 |
| 2 | 6,041 | 367 | 6,408 | 360,924 |
| 3 | 6,047 | 361 | 6,408 | 354,876 |
| 4 | 6,053 | 355 | 6,408 | 348,823 |
| 5 | 6,059 | 349 | 6,408 | 342,764 |
| 6 | 6,065 | 343 | 6,408 | 336,699 |
| 7 | 6,071 | 337 | 6,408 | 330,627 |
| 8 | 6,078 | 331 | 6,408 | 324,550 |
| 9 | 6,084 | 325 | 6,408 | 318,466 |
| 10 | 6,090 | 318 | 6,408 | 312,376 |
| 11 | 6,096 | 312 | 6,408 | 306,281 |
| 12 | 6,102 | 306 | 6,408 | 300,179 |
| 13 | 6,108 | 300 | 6,408 | 294,071 |
| 14 | 6,114 | 294 | 6,408 | 287,957 |
| 15 | 6,120 | 288 | 6,408 | 281,836 |
| 16 | 6,126 | 282 | 6,408 | 275,710 |
| 17 | 6,132 | 276 | 6,408 | 269,578 |
| 18 | 6,139 | 270 | 6,408 | 263,439 |
| 19 | 6,145 | 263 | 6,408 | 257,294 |
| 20 | 6,151 | 257 | 6,408 | 251,144 |
| 21 | 6,157 | 251 | 6,408 | 244,987 |
| 22 | 6,163 | 245 | 6,408 | 238,823 |
| 23 | 6,169 | 239 | 6,408 | 232,654 |
| 24 | 6,175 | 233 | 6,408 | 226,479 |
| 25 | 6,182 | 226 | 6,408 | 220,297 |
| 26 | 6,188 | 220 | 6,408 | 214,109 |
| 27 | 6,194 | 214 | 6,408 | 207,915 |
| 28 | 6,200 | 208 | 6,408 | 201,715 |
| 29 | 6,206 | 202 | 6,408 | 195,509 |
| 30 | 6,213 | 196 | 6,408 | 189,296 |
| 31 | 6,219 | 189 | 6,408 | 183,077 |
| 32 | 6,225 | 183 | 6,408 | 176,852 |
| 33 | 6,231 | 177 | 6,408 | 170,621 |
| 34 | 6,238 | 171 | 6,408 | 164,383 |
| 35 | 6,244 | 164 | 6,408 | 158,139 |
| 36 | 6,250 | 158 | 6,408 | 151,889 |
| 37 | 6,256 | 152 | 6,408 | 145,633 |
| 38 | 6,263 | 146 | 6,408 | 139,371 |
| 39 | 6,269 | 139 | 6,408 | 133,102 |
| 40 | 6,275 | 133 | 6,408 | 126,827 |
| 41 | 6,281 | 127 | 6,408 | 120,546 |
| 42 | 6,288 | 121 | 6,408 | 114,258 |
| 43 | 6,294 | 114 | 6,408 | 107,964 |
| 44 | 6,300 | 108 | 6,408 | 101,664 |
| 45 | 6,306 | 102 | 6,408 | 95,357 |
| 46 | 6,313 | 95 | 6,408 | 89,045 |
| 47 | 6,319 | 89 | 6,408 | 82,726 |
| 48 | 6,325 | 83 | 6,408 | 76,400 |
| 49 | 6,332 | 76 | 6,408 | 70,068 |
| 50 | 6,338 | 70 | 6,408 | 63,730 |
| 51 | 6,344 | 64 | 6,408 | 57,386 |
| 52 | 6,351 | 57 | 6,408 | 51,035 |
| 53 | 6,357 | 51 | 6,408 | 44,678 |
| 54 | 6,363 | 45 | 6,408 | 38,315 |
| 55 | 6,370 | 38 | 6,408 | 31,945 |
| 56 | 6,376 | 32 | 6,408 | 25,569 |
| 57 | 6,383 | 26 | 6,408 | 19,186 |
| 58 | 6,389 | 19 | 6,408 | 12,797 |
| 59 | 6,395 | 13 | 6,408 | 6,402 |
| 60 | 6,402 | 6 | 6,408 | 0 |

#### Supplementary 21

Straight-Line GRT Amortization Table of axicabtagene ciloleucel (Yescarta®) at an interest rate of 1% monthly over 10 years

| **Payment Period** | **Principal Amount Due** | **Interest Amount Due** | **Total Payment Amount Due** | **Principal Balance** |
| --- | --- | --- | --- | --- |
|  |  |  |  | 373,000 |
| 1 | 2,927 | 373 | 3,300 | 370,073 |
| 2 | 2,930 | 370 | 3,300 | 367,143 |
| 3 | 2,933 | 367 | 3,300 | 364,210 |
| 4 | 2,936 | 364 | 3,300 | 361,274 |
| 5 | 2,939 | 361 | 3,300 | 358,335 |
| 6 | 2,942 | 358 | 3,300 | 355,393 |
| 7 | 2,945 | 355 | 3,300 | 352,449 |
| 8 | 2,948 | 352 | 3,300 | 349,501 |
| 9 | 2,951 | 350 | 3,300 | 346,550 |
| 10 | 2,954 | 347 | 3,300 | 343,597 |
| 11 | 2,957 | 344 | 3,300 | 340,640 |
| 12 | 2,959 | 341 | 3,300 | 337,681 |
| 13 | 2,962 | 338 | 3,300 | 334,718 |
| 14 | 2,965 | 335 | 3,300 | 331,753 |
| 15 | 2,968 | 332 | 3,300 | 328,785 |
| 16 | 2,971 | 329 | 3,300 | 325,813 |
| 17 | 2,974 | 326 | 3,300 | 322,839 |
| 18 | 2,977 | 323 | 3,300 | 319,862 |
| 19 | 2,980 | 320 | 3,300 | 316,881 |
| 20 | 2,983 | 317 | 3,300 | 313,898 |
| 21 | 2,986 | 314 | 3,300 | 310,912 |
| 22 | 2,989 | 311 | 3,300 | 307,923 |
| 23 | 2,992 | 308 | 3,300 | 304,931 |
| 24 | 2,995 | 305 | 3,300 | 301,935 |
| 25 | 2,998 | 302 | 3,300 | 298,937 |
| 26 | 3,001 | 299 | 3,300 | 295,936 |
| 27 | 3,004 | 296 | 3,300 | 292,932 |
| 28 | 3,007 | 293 | 3,300 | 289,925 |
| 29 | 3,010 | 290 | 3,300 | 286,915 |
| 30 | 3,013 | 287 | 3,300 | 283,901 |
| 31 | 3,016 | 284 | 3,300 | 280,885 |
| 32 | 3,019 | 281 | 3,300 | 277,866 |
| 33 | 3,022 | 278 | 3,300 | 274,844 |
| 34 | 3,025 | 275 | 3,300 | 271,818 |
| 35 | 3,028 | 272 | 3,300 | 268,790 |
| 36 | 3,031 | 269 | 3,300 | 265,759 |
| 37 | 3,034 | 266 | 3,300 | 262,724 |
| 38 | 3,037 | 263 | 3,300 | 259,687 |
| 39 | 3,040 | 260 | 3,300 | 256,647 |
| 40 | 3,043 | 257 | 3,300 | 253,603 |
| 41 | 3,047 | 254 | 3,300 | 250,557 |
| 42 | 3,050 | 251 | 3,300 | 247,507 |
| 43 | 3,053 | 248 | 3,300 | 244,454 |
| 44 | 3,056 | 244 | 3,300 | 241,399 |
| 45 | 3,059 | 241 | 3,300 | 238,340 |
| 46 | 3,062 | 238 | 3,300 | 235,278 |
| 47 | 3,065 | 235 | 3,300 | 232,213 |
| 48 | 3,068 | 232 | 3,300 | 229,146 |
| 49 | 3,071 | 229 | 3,300 | 226,075 |
| 50 | 3,074 | 226 | 3,300 | 223,001 |
| 51 | 3,077 | 223 | 3,300 | 219,923 |
| 52 | 3,080 | 220 | 3,300 | 216,843 |
| 53 | 3,083 | 217 | 3,300 | 213,760 |
| 54 | 3,086 | 214 | 3,300 | 210,674 |
| 55 | 3,089 | 211 | 3,300 | 207,584 |
| 56 | 3,093 | 208 | 3,300 | 204,492 |
| 57 | 3,096 | 204 | 3,300 | 201,396 |
| 58 | 3,099 | 201 | 3,300 | 198,297 |
| 59 | 3,102 | 198 | 3,300 | 195,195 |
| 60 | 3,105 | 195 | 3,300 | 192,091 |
| 61 | 3,108 | 192 | 3,300 | 188,983 |
| 62 | 3,111 | 189 | 3,300 | 185,871 |
| 63 | 3,114 | 186 | 3,300 | 182,757 |
| 64 | 3,117 | 183 | 3,300 | 179,640 |
| 65 | 3,120 | 180 | 3,300 | 176,519 |
| 66 | 3,124 | 177 | 3,300 | 173,396 |
| 67 | 3,127 | 173 | 3,300 | 170,269 |
| 68 | 3,130 | 170 | 3,300 | 167,139 |
| 69 | 3,133 | 167 | 3,300 | 164,006 |
| 70 | 3,136 | 164 | 3,300 | 160,870 |
| 71 | 3,139 | 161 | 3,300 | 157,731 |
| 72 | 3,142 | 158 | 3,300 | 154,588 |
| 73 | 3,146 | 155 | 3,300 | 151,443 |
| 74 | 3,149 | 151 | 3,300 | 148,294 |
| 75 | 3,152 | 148 | 3,300 | 145,142 |
| 76 | 3,155 | 145 | 3,300 | 141,987 |
| 77 | 3,158 | 142 | 3,300 | 138,829 |
| 78 | 3,161 | 139 | 3,300 | 135,668 |
| 79 | 3,164 | 136 | 3,300 | 132,504 |
| 80 | 3,168 | 133 | 3,300 | 129,336 |
| 81 | 3,171 | 129 | 3,300 | 126,165 |
| 82 | 3,174 | 126 | 3,300 | 122,991 |
| 83 | 3,177 | 123 | 3,300 | 119,814 |
| 84 | 3,180 | 120 | 3,300 | 116,634 |
| 85 | 3,183 | 117 | 3,300 | 113,450 |
| 86 | 3,187 | 113 | 3,300 | 110,264 |
| 87 | 3,190 | 110 | 3,300 | 107,074 |
| 88 | 3,193 | 107 | 3,300 | 103,881 |
| 89 | 3,196 | 104 | 3,300 | 100,685 |
| 90 | 3,199 | 101 | 3,300 | 97,485 |
| 91 | 3,203 | 97 | 3,300 | 94,282 |
| 92 | 3,206 | 94 | 3,300 | 91,077 |
| 93 | 3,209 | 91 | 3,300 | 87,868 |
| 94 | 3,212 | 88 | 3,300 | 84,655 |
| 95 | 3,215 | 85 | 3,300 | 81,440 |
| 96 | 3,219 | 81 | 3,300 | 78,221 |
| 97 | 3,222 | 78 | 3,300 | 74,999 |
| 98 | 3,225 | 75 | 3,300 | 71,774 |
| 99 | 3,228 | 72 | 3,300 | 68,546 |
| 100 | 3,232 | 69 | 3,300 | 65,314 |
| 101 | 3,235 | 65 | 3,300 | 62,080 |
| 102 | 3,238 | 62 | 3,300 | 58,841 |
| 103 | 3,241 | 59 | 3,300 | 55,600 |
| 104 | 3,245 | 56 | 3,300 | 52,356 |
| 105 | 3,248 | 52 | 3,300 | 49,108 |
| 106 | 3,251 | 49 | 3,300 | 45,857 |
| 107 | 3,254 | 46 | 3,300 | 42,603 |
| 108 | 3,258 | 43 | 3,300 | 39,345 |
| 109 | 3,261 | 39 | 3,300 | 36,084 |
| 110 | 3,264 | 36 | 3,300 | 32,820 |
| 111 | 3,267 | 33 | 3,300 | 29,553 |
| 112 | 3,271 | 30 | 3,300 | 26,283 |
| 113 | 3,274 | 26 | 3,300 | 23,009 |
| 114 | 3,277 | 23 | 3,300 | 19,732 |
| 115 | 3,280 | 20 | 3,300 | 16,451 |
| 116 | 3,284 | 16 | 3,300 | 13,168 |
| 117 | 3,287 | 13 | 3,300 | 9,881 |
| 118 | 3,290 | 10 | 3,300 | 6,590 |
| 119 | 3,294 | 7 | 3,300 | 3,297 |
| 120 | 3,297 | 3 | 3,300 | 0 |

#### Supplementary 22

Straight-Line GRT Amortization Table of axicabtagene ciloleucel (Yescarta®) at an interest rate of 3% monthly over 5 years

| **Payment Period** | **Principal Amount Due** | **Interest Amount Due** | **Total Payment Amount Due** | **Principal Balance** |
| --- | --- | --- | --- | --- |
|  |  |  |  | 373,000 |
| 1 | 5,683 | 1,119 | 6,802 | 367,317 |
| 2 | 5,700 | 1,102 | 6,802 | 361,616 |
| 3 | 5,717 | 1,085 | 6,802 | 355,899 |
| 4 | 5,735 | 1,068 | 6,802 | 350,165 |
| 5 | 5,752 | 1,050 | 6,802 | 344,413 |
| 6 | 5,769 | 1,033 | 6,802 | 338,644 |
| 7 | 5,786 | 1,016 | 6,802 | 332,857 |
| 8 | 5,804 | 999 | 6,802 | 327,054 |
| 9 | 5,821 | 981 | 6,802 | 321,233 |
| 10 | 5,839 | 964 | 6,802 | 315,394 |
| 11 | 5,856 | 946 | 6,802 | 309,538 |
| 12 | 5,874 | 929 | 6,802 | 303,665 |
| 13 | 5,891 | 911 | 6,802 | 297,773 |
| 14 | 5,909 | 893 | 6,802 | 291,864 |
| 15 | 5,927 | 876 | 6,802 | 285,938 |
| 16 | 5,944 | 858 | 6,802 | 279,993 |
| 17 | 5,962 | 840 | 6,802 | 274,031 |
| 18 | 5,980 | 822 | 6,802 | 268,051 |
| 19 | 5,998 | 804 | 6,802 | 262,053 |
| 20 | 6,016 | 786 | 6,802 | 256,037 |
| 21 | 6,034 | 768 | 6,802 | 250,003 |
| 22 | 6,052 | 750 | 6,802 | 243,950 |
| 23 | 6,070 | 732 | 6,802 | 237,880 |
| 24 | 6,089 | 714 | 6,802 | 231,791 |
| 25 | 6,107 | 695 | 6,802 | 225,685 |
| 26 | 6,125 | 677 | 6,802 | 219,559 |
| 27 | 6,144 | 659 | 6,802 | 213,416 |
| 28 | 6,162 | 640 | 6,802 | 207,254 |
| 29 | 6,180 | 622 | 6,802 | 201,073 |
| 30 | 6,199 | 603 | 6,802 | 194,874 |
| 31 | 6,218 | 585 | 6,802 | 188,657 |
| 32 | 6,236 | 566 | 6,802 | 182,420 |
| 33 | 6,255 | 547 | 6,802 | 176,165 |
| 34 | 6,274 | 528 | 6,802 | 169,892 |
| 35 | 6,293 | 510 | 6,802 | 163,599 |
| 36 | 6,311 | 491 | 6,802 | 157,288 |
| 37 | 6,330 | 472 | 6,802 | 150,957 |
| 38 | 6,349 | 453 | 6,802 | 144,608 |
| 39 | 6,368 | 434 | 6,802 | 138,240 |
| 40 | 6,388 | 415 | 6,802 | 131,852 |
| 41 | 6,407 | 396 | 6,802 | 125,445 |
| 42 | 6,426 | 376 | 6,802 | 119,019 |
| 43 | 6,445 | 357 | 6,802 | 112,574 |
| 44 | 6,465 | 338 | 6,802 | 106,110 |
| 45 | 6,484 | 318 | 6,802 | 99,626 |
| 46 | 6,503 | 299 | 6,802 | 93,122 |
| 47 | 6,523 | 279 | 6,802 | 86,600 |
| 48 | 6,542 | 260 | 6,802 | 80,057 |
| 49 | 6,562 | 240 | 6,802 | 73,495 |
| 50 | 6,582 | 220 | 6,802 | 66,913 |
| 51 | 6,601 | 201 | 6,802 | 60,312 |
| 52 | 6,621 | 181 | 6,802 | 53,691 |
| 53 | 6,641 | 161 | 6,802 | 47,049 |
| 54 | 6,661 | 141 | 6,802 | 40,388 |
| 55 | 6,681 | 121 | 6,802 | 33,707 |
| 56 | 6,701 | 101 | 6,802 | 27,006 |
| 57 | 6,721 | 81 | 6,802 | 20,285 |
| 58 | 6,741 | 61 | 6,802 | 13,543 |
| 59 | 6,762 | 41 | 6,802 | 6,782 |
| 60 | 6,782 | 20 | 6,802 | 0 |

#### Supplementary 23

Straight-Line GRT Amortization Table of axicabtagene ciloleucel (Yescarta®) at an interest rate of 3% monthly over 10 years

| **Payment Period** | **Principal Amount Due** | **Interest Amount Due** | **Total Payment Amount Due** | **Principal Balance** |
| --- | --- | --- | --- | --- |
|  |  |  |  | 373,000 |
| 1 | 2,587 | 1,119 | 3,706 | 370,413 |
| 2 | 2,595 | 1,111 | 3,706 | 367,818 |
| 3 | 2,602 | 1,103 | 3,706 | 365,216 |
| 4 | 2,610 | 1,096 | 3,706 | 362,606 |
| 5 | 2,618 | 1,088 | 3,706 | 359,987 |
| 6 | 2,626 | 1,080 | 3,706 | 357,361 |
| 7 | 2,634 | 1,072 | 3,706 | 354,728 |
| 8 | 2,642 | 1,064 | 3,706 | 352,086 |
| 9 | 2,650 | 1,056 | 3,706 | 349,436 |
| 10 | 2,658 | 1,048 | 3,706 | 346,779 |
| 11 | 2,666 | 1,040 | 3,706 | 344,113 |
| 12 | 2,674 | 1,032 | 3,706 | 341,439 |
| 13 | 2,682 | 1,024 | 3,706 | 338,758 |
| 14 | 2,690 | 1,016 | 3,706 | 336,068 |
| 15 | 2,698 | 1,008 | 3,706 | 333,370 |
| 16 | 2,706 | 1,000 | 3,706 | 330,664 |
| 17 | 2,714 | 992 | 3,706 | 327,951 |
| 18 | 2,722 | 984 | 3,706 | 325,228 |
| 19 | 2,730 | 976 | 3,706 | 322,498 |
| 20 | 2,738 | 967 | 3,706 | 319,760 |
| 21 | 2,747 | 959 | 3,706 | 317,013 |
| 22 | 2,755 | 951 | 3,706 | 314,258 |
| 23 | 2,763 | 943 | 3,706 | 311,495 |
| 24 | 2,771 | 934 | 3,706 | 308,724 |
| 25 | 2,780 | 926 | 3,706 | 305,944 |
| 26 | 2,788 | 918 | 3,706 | 303,156 |
| 27 | 2,796 | 909 | 3,706 | 300,359 |
| 28 | 2,805 | 901 | 3,706 | 297,554 |
| 29 | 2,813 | 893 | 3,706 | 294,741 |
| 30 | 2,822 | 884 | 3,706 | 291,919 |
| 31 | 2,830 | 876 | 3,706 | 289,089 |
| 32 | 2,839 | 867 | 3,706 | 286,250 |
| 33 | 2,847 | 859 | 3,706 | 283,403 |
| 34 | 2,856 | 850 | 3,706 | 280,548 |
| 35 | 2,864 | 842 | 3,706 | 277,683 |
| 36 | 2,873 | 833 | 3,706 | 274,810 |
| 37 | 2,882 | 824 | 3,706 | 271,929 |
| 38 | 2,890 | 816 | 3,706 | 269,039 |
| 39 | 2,899 | 807 | 3,706 | 266,140 |
| 40 | 2,908 | 798 | 3,706 | 263,232 |
| 41 | 2,916 | 790 | 3,706 | 260,316 |
| 42 | 2,925 | 781 | 3,706 | 257,391 |
| 43 | 2,934 | 772 | 3,706 | 254,457 |
| 44 | 2,943 | 763 | 3,706 | 251,515 |
| 45 | 2,951 | 755 | 3,706 | 248,563 |
| 46 | 2,960 | 746 | 3,706 | 245,603 |
| 47 | 2,969 | 737 | 3,706 | 242,634 |
| 48 | 2,978 | 728 | 3,706 | 239,656 |
| 49 | 2,987 | 719 | 3,706 | 236,669 |
| 50 | 2,996 | 710 | 3,706 | 233,673 |
| 51 | 3,005 | 701 | 3,706 | 230,668 |
| 52 | 3,014 | 692 | 3,706 | 227,654 |
| 53 | 3,023 | 683 | 3,706 | 224,631 |
| 54 | 3,032 | 674 | 3,706 | 221,599 |
| 55 | 3,041 | 665 | 3,706 | 218,558 |
| 56 | 3,050 | 656 | 3,706 | 215,508 |
| 57 | 3,059 | 647 | 3,706 | 212,448 |
| 58 | 3,069 | 637 | 3,706 | 209,380 |
| 59 | 3,078 | 628 | 3,706 | 206,302 |
| 60 | 3,087 | 619 | 3,706 | 203,215 |
| 61 | 3,096 | 610 | 3,706 | 200,119 |
| 62 | 3,106 | 600 | 3,706 | 197,013 |
| 63 | 3,115 | 591 | 3,706 | 193,898 |
| 64 | 3,124 | 582 | 3,706 | 190,774 |
| 65 | 3,134 | 572 | 3,706 | 187,640 |
| 66 | 3,143 | 563 | 3,706 | 184,497 |
| 67 | 3,152 | 553 | 3,706 | 181,345 |
| 68 | 3,162 | 544 | 3,706 | 178,183 |
| 69 | 3,171 | 535 | 3,706 | 175,011 |
| 70 | 3,181 | 525 | 3,706 | 171,831 |
| 71 | 3,190 | 515 | 3,706 | 168,640 |
| 72 | 3,200 | 506 | 3,706 | 165,440 |
| 73 | 3,210 | 496 | 3,706 | 162,230 |
| 74 | 3,219 | 487 | 3,706 | 159,011 |
| 75 | 3,229 | 477 | 3,706 | 155,782 |
| 76 | 3,239 | 467 | 3,706 | 152,544 |
| 77 | 3,248 | 458 | 3,706 | 149,295 |
| 78 | 3,258 | 448 | 3,706 | 146,037 |
| 79 | 3,268 | 438 | 3,706 | 142,770 |
| 80 | 3,278 | 428 | 3,706 | 139,492 |
| 81 | 3,287 | 418 | 3,706 | 136,204 |
| 82 | 3,297 | 409 | 3,706 | 132,907 |
| 83 | 3,307 | 399 | 3,706 | 129,600 |
| 84 | 3,317 | 389 | 3,706 | 126,283 |
| 85 | 3,327 | 379 | 3,706 | 122,956 |
| 86 | 3,337 | 369 | 3,706 | 119,619 |
| 87 | 3,347 | 359 | 3,706 | 116,271 |
| 88 | 3,357 | 349 | 3,706 | 112,914 |
| 89 | 3,367 | 339 | 3,706 | 109,547 |
| 90 | 3,377 | 329 | 3,706 | 106,170 |
| 91 | 3,387 | 319 | 3,706 | 102,782 |
| 92 | 3,398 | 308 | 3,706 | 99,385 |
| 93 | 3,408 | 298 | 3,706 | 95,977 |
| 94 | 3,418 | 288 | 3,706 | 92,559 |
| 95 | 3,428 | 278 | 3,706 | 89,131 |
| 96 | 3,439 | 267 | 3,706 | 85,692 |
| 97 | 3,449 | 257 | 3,706 | 82,243 |
| 98 | 3,459 | 247 | 3,706 | 78,784 |
| 99 | 3,470 | 236 | 3,706 | 75,315 |
| 100 | 3,480 | 226 | 3,706 | 71,835 |
| 101 | 3,490 | 216 | 3,706 | 68,344 |
| 102 | 3,501 | 205 | 3,706 | 64,843 |
| 103 | 3,511 | 195 | 3,706 | 61,332 |
| 104 | 3,522 | 184 | 3,706 | 57,810 |
| 105 | 3,533 | 173 | 3,706 | 54,277 |
| 106 | 3,543 | 163 | 3,706 | 50,734 |
| 107 | 3,554 | 152 | 3,706 | 47,181 |
| 108 | 3,564 | 142 | 3,706 | 43,616 |
| 109 | 3,575 | 131 | 3,706 | 40,041 |
| 110 | 3,586 | 120 | 3,706 | 36,455 |
| 111 | 3,597 | 109 | 3,706 | 32,859 |
| 112 | 3,607 | 99 | 3,706 | 29,251 |
| 113 | 3,618 | 88 | 3,706 | 25,633 |
| 114 | 3,629 | 77 | 3,706 | 22,004 |
| 115 | 3,640 | 66 | 3,706 | 18,364 |
| 116 | 3,651 | 55 | 3,706 | 14,713 |
| 117 | 3,662 | 44 | 3,706 | 11,051 |
| 118 | 3,673 | 33 | 3,706 | 7,379 |
| 119 | 3,684 | 22 | 3,706 | 3,695 |
| 120 | 3,695 | 11 | 3,706 | 0 |

#### Supplementary 24

Straight-Line GRT Amortization Table of tisagenlecleucel (Kymriah®) at an interest rate of 1% monthly over 5 years

| **Payment Period** | **Principal Amount Due** | **Interest Amount Due** | **Total Payment Amount Due** | **Principal Balance** |
| --- | --- | --- | --- | --- |
|  |  |  |  | 475,000 |
| 1 | 7,685 | 475 | 8,160 | 467,315 |
| 2 | 7,693 | 467 | 8,160 | 459,621 |
| 3 | 7,701 | 460 | 8,160 | 451,920 |
| 4 | 7,709 | 452 | 8,160 | 444,212 |
| 5 | 7,716 | 444 | 8,160 | 436,496 |
| 6 | 7,724 | 436 | 8,160 | 428,772 |
| 7 | 7,732 | 429 | 8,160 | 421,040 |
| 8 | 7,739 | 421 | 8,160 | 413,300 |
| 9 | 7,747 | 413 | 8,160 | 405,553 |
| 10 | 7,755 | 406 | 8,160 | 397,798 |
| 11 | 7,763 | 398 | 8,160 | 390,036 |
| 12 | 7,770 | 390 | 8,160 | 382,265 |
| 13 | 7,778 | 382 | 8,160 | 374,487 |
| 14 | 7,786 | 374 | 8,160 | 366,701 |
| 15 | 7,794 | 367 | 8,160 | 358,907 |
| 16 | 7,802 | 359 | 8,160 | 351,105 |
| 17 | 7,809 | 351 | 8,160 | 343,296 |
| 18 | 7,817 | 343 | 8,160 | 335,479 |
| 19 | 7,825 | 335 | 8,160 | 327,654 |
| 20 | 7,833 | 328 | 8,160 | 319,821 |
| 21 | 7,841 | 320 | 8,160 | 311,980 |
| 22 | 7,849 | 312 | 8,160 | 304,132 |
| 23 | 7,856 | 304 | 8,160 | 296,275 |
| 24 | 7,864 | 296 | 8,160 | 288,411 |
| 25 | 7,872 | 288 | 8,160 | 280,539 |
| 26 | 7,880 | 281 | 8,160 | 272,659 |
| 27 | 7,888 | 273 | 8,160 | 264,771 |
| 28 | 7,896 | 265 | 8,160 | 256,876 |
| 29 | 7,904 | 257 | 8,160 | 248,972 |
| 30 | 7,912 | 249 | 8,160 | 241,060 |
| 31 | 7,919 | 241 | 8,160 | 233,141 |
| 32 | 7,927 | 233 | 8,160 | 225,214 |
| 33 | 7,935 | 225 | 8,160 | 217,278 |
| 34 | 7,943 | 217 | 8,160 | 209,335 |
| 35 | 7,951 | 209 | 8,160 | 201,384 |
| 36 | 7,959 | 201 | 8,160 | 193,425 |
| 37 | 7,967 | 193 | 8,160 | 185,458 |
| 38 | 7,975 | 185 | 8,160 | 177,483 |
| 39 | 7,983 | 177 | 8,160 | 169,500 |
| 40 | 7,991 | 169 | 8,160 | 161,509 |
| 41 | 7,999 | 162 | 8,160 | 153,510 |
| 42 | 8,007 | 154 | 8,160 | 145,503 |
| 43 | 8,015 | 146 | 8,160 | 137,488 |
| 44 | 8,023 | 137 | 8,160 | 129,465 |
| 45 | 8,031 | 129 | 8,160 | 121,434 |
| 46 | 8,039 | 121 | 8,160 | 113,395 |
| 47 | 8,047 | 113 | 8,160 | 105,348 |
| 48 | 8,055 | 105 | 8,160 | 97,292 |
| 49 | 8,063 | 97 | 8,160 | 89,229 |
| 50 | 8,071 | 89 | 8,160 | 81,158 |
| 51 | 8,079 | 81 | 8,160 | 73,079 |
| 52 | 8,087 | 73 | 8,160 | 64,991 |
| 53 | 8,096 | 65 | 8,160 | 56,896 |
| 54 | 8,104 | 57 | 8,160 | 48,792 |
| 55 | 8,112 | 49 | 8,160 | 40,680 |
| 56 | 8,120 | 41 | 8,160 | 32,561 |
| 57 | 8,128 | 33 | 8,160 | 24,433 |
| 58 | 8,136 | 24 | 8,160 | 16,297 |
| 59 | 8,144 | 16 | 8,160 | 8,152 |
| 60 | 8,152 | 8 | 8,160 | 0 |

#### Supplementary 25

Straight-Line GRT Amortization Table of tisagenlecleucel (Kymriah®) at an interest rate of 1% monthly over 10 years

| **Payment Period** | **Principal Amount Due** | **Interest Amount Due** | **Total Payment Amount Due** | **Principal Balance** |
| --- | --- | --- | --- | --- |
|  |  |  |  | 475,000 |
| 1 | 3,728 | 475 | 4,203 | 471,272 |
| 2 | 3,731 | 471 | 4,203 | 467,541 |
| 3 | 3,735 | 468 | 4,203 | 463,806 |
| 4 | 3,739 | 464 | 4,203 | 460,067 |
| 5 | 3,742 | 460 | 4,203 | 456,325 |
| 6 | 3,746 | 456 | 4,203 | 452,579 |
| 7 | 3,750 | 453 | 4,203 | 448,829 |
| 8 | 3,754 | 449 | 4,203 | 445,075 |
| 9 | 3,757 | 445 | 4,203 | 441,317 |
| 10 | 3,761 | 441 | 4,203 | 437,556 |
| 11 | 3,765 | 438 | 4,203 | 433,791 |
| 12 | 3,769 | 434 | 4,203 | 430,022 |
| 13 | 3,773 | 430 | 4,203 | 426,250 |
| 14 | 3,776 | 426 | 4,203 | 422,474 |
| 15 | 3,780 | 422 | 4,203 | 418,694 |
| 16 | 3,784 | 419 | 4,203 | 414,910 |
| 17 | 3,788 | 415 | 4,203 | 411,122 |
| 18 | 3,791 | 411 | 4,203 | 407,331 |
| 19 | 3,795 | 407 | 4,203 | 403,535 |
| 20 | 3,799 | 404 | 4,203 | 399,736 |
| 21 | 3,803 | 400 | 4,203 | 395,934 |
| 22 | 3,807 | 396 | 4,203 | 392,127 |
| 23 | 3,810 | 392 | 4,203 | 388,316 |
| 24 | 3,814 | 388 | 4,203 | 384,502 |
| 25 | 3,818 | 385 | 4,203 | 380,684 |
| 26 | 3,822 | 381 | 4,203 | 376,862 |
| 27 | 3,826 | 377 | 4,203 | 373,037 |
| 28 | 3,830 | 373 | 4,203 | 369,207 |
| 29 | 3,833 | 369 | 4,203 | 365,374 |
| 30 | 3,837 | 365 | 4,203 | 361,537 |
| 31 | 3,841 | 362 | 4,203 | 357,695 |
| 32 | 3,845 | 358 | 4,203 | 353,851 |
| 33 | 3,849 | 354 | 4,203 | 350,002 |
| 34 | 3,853 | 350 | 4,203 | 346,149 |
| 35 | 3,856 | 346 | 4,203 | 342,293 |
| 36 | 3,860 | 342 | 4,203 | 338,433 |
| 37 | 3,864 | 338 | 4,203 | 334,569 |
| 38 | 3,868 | 335 | 4,203 | 330,701 |
| 39 | 3,872 | 331 | 4,203 | 326,829 |
| 40 | 3,876 | 327 | 4,203 | 322,953 |
| 41 | 3,880 | 323 | 4,203 | 319,073 |
| 42 | 3,883 | 319 | 4,203 | 315,190 |
| 43 | 3,887 | 315 | 4,203 | 311,303 |
| 44 | 3,891 | 311 | 4,203 | 307,411 |
| 45 | 3,895 | 307 | 4,203 | 303,516 |
| 46 | 3,899 | 304 | 4,203 | 299,617 |
| 47 | 3,903 | 300 | 4,203 | 295,714 |
| 48 | 3,907 | 296 | 4,203 | 291,807 |
| 49 | 3,911 | 292 | 4,203 | 287,897 |
| 50 | 3,915 | 288 | 4,203 | 283,982 |
| 51 | 3,919 | 284 | 4,203 | 280,063 |
| 52 | 3,922 | 280 | 4,203 | 276,141 |
| 53 | 3,926 | 276 | 4,203 | 272,214 |
| 54 | 3,930 | 272 | 4,203 | 268,284 |
| 55 | 3,934 | 268 | 4,203 | 264,350 |
| 56 | 3,938 | 264 | 4,203 | 260,412 |
| 57 | 3,942 | 260 | 4,203 | 256,469 |
| 58 | 3,946 | 256 | 4,203 | 252,523 |
| 59 | 3,950 | 253 | 4,203 | 248,573 |
| 60 | 3,954 | 249 | 4,203 | 244,619 |
| 61 | 3,958 | 245 | 4,203 | 240,661 |
| 62 | 3,962 | 241 | 4,203 | 236,699 |
| 63 | 3,966 | 237 | 4,203 | 232,734 |
| 64 | 3,970 | 233 | 4,203 | 228,764 |
| 65 | 3,974 | 229 | 4,203 | 224,790 |
| 66 | 3,978 | 225 | 4,203 | 220,812 |
| 67 | 3,982 | 221 | 4,203 | 216,830 |
| 68 | 3,986 | 217 | 4,203 | 212,845 |
| 69 | 3,990 | 213 | 4,203 | 208,855 |
| 70 | 3,994 | 209 | 4,203 | 204,861 |
| 71 | 3,998 | 205 | 4,203 | 200,864 |
| 72 | 4,002 | 201 | 4,203 | 196,862 |
| 73 | 4,006 | 197 | 4,203 | 192,856 |
| 74 | 4,010 | 193 | 4,203 | 188,847 |
| 75 | 4,014 | 189 | 4,203 | 184,833 |
| 76 | 4,018 | 185 | 4,203 | 180,815 |
| 77 | 4,022 | 181 | 4,203 | 176,793 |
| 78 | 4,026 | 177 | 4,203 | 172,768 |
| 79 | 4,030 | 173 | 4,203 | 168,738 |
| 80 | 4,034 | 169 | 4,203 | 164,704 |
| 81 | 4,038 | 165 | 4,203 | 160,666 |
| 82 | 4,042 | 161 | 4,203 | 156,624 |
| 83 | 4,046 | 157 | 4,203 | 152,578 |
| 84 | 4,050 | 153 | 4,203 | 148,528 |
| 85 | 4,054 | 149 | 4,203 | 144,474 |
| 86 | 4,058 | 144 | 4,203 | 140,416 |
| 87 | 4,062 | 140 | 4,203 | 136,354 |
| 88 | 4,066 | 136 | 4,203 | 132,288 |
| 89 | 4,070 | 132 | 4,203 | 128,218 |
| 90 | 4,074 | 128 | 4,203 | 124,143 |
| 91 | 4,078 | 124 | 4,203 | 120,065 |
| 92 | 4,082 | 120 | 4,203 | 115,982 |
| 93 | 4,087 | 116 | 4,203 | 111,896 |
| 94 | 4,091 | 112 | 4,203 | 107,805 |
| 95 | 4,095 | 108 | 4,203 | 103,710 |
| 96 | 4,099 | 104 | 4,203 | 99,611 |
| 97 | 4,103 | 100 | 4,203 | 95,509 |
| 98 | 4,107 | 96 | 4,203 | 91,401 |
| 99 | 4,111 | 91 | 4,203 | 87,290 |
| 100 | 4,115 | 87 | 4,203 | 83,175 |
| 101 | 4,119 | 83 | 4,203 | 79,056 |
| 102 | 4,124 | 79 | 4,203 | 74,932 |
| 103 | 4,128 | 75 | 4,203 | 70,805 |
| 104 | 4,132 | 71 | 4,203 | 66,673 |
| 105 | 4,136 | 67 | 4,203 | 62,537 |
| 106 | 4,140 | 63 | 4,203 | 58,397 |
| 107 | 4,144 | 58 | 4,203 | 54,253 |
| 108 | 4,148 | 54 | 4,203 | 50,104 |
| 109 | 4,152 | 50 | 4,203 | 45,952 |
| 110 | 4,157 | 46 | 4,203 | 41,795 |
| 111 | 4,161 | 42 | 4,203 | 37,635 |
| 112 | 4,165 | 38 | 4,203 | 33,470 |
| 113 | 4,169 | 33 | 4,203 | 29,301 |
| 114 | 4,173 | 29 | 4,203 | 25,127 |
| 115 | 4,177 | 25 | 4,203 | 20,950 |
| 116 | 4,182 | 21 | 4,203 | 16,768 |
| 117 | 4,186 | 17 | 4,203 | 12,583 |
| 118 | 4,190 | 13 | 4,203 | 8,393 |
| 119 | 4,194 | 8 | 4,203 | 4,198 |
| 120 | 4,198 | 4 | 4,203 | 0 |

#### Supplementary 26

Straight-Line GRT Amortization Table of tisagenlecleucel (Kymriah®) at an interest rate of 3% monthly over 5 years

| **Payment Period** | **Principal Amount Due** | **Interest Amount Due** | **Total Payment Amount Due** | **Principal Balance** |
| --- | --- | --- | --- | --- |
|  |  |  |  | 475,000 |
| 1 | 7,237 | 1,425 | 8,662 | 467,763 |
| 2 | 7,259 | 1,403 | 8,662 | 460,504 |
| 3 | 7,281 | 1,382 | 8,662 | 453,223 |
| 4 | 7,303 | 1,360 | 8,662 | 445,920 |
| 5 | 7,325 | 1,338 | 8,662 | 438,595 |
| 6 | 7,347 | 1,316 | 8,662 | 431,249 |
| 7 | 7,369 | 1,294 | 8,662 | 423,880 |
| 8 | 7,391 | 1,272 | 8,662 | 416,489 |
| 9 | 7,413 | 1,249 | 8,662 | 409,077 |
| 10 | 7,435 | 1,227 | 8,662 | 401,641 |
| 11 | 7,457 | 1,205 | 8,662 | 394,184 |
| 12 | 7,480 | 1,183 | 8,662 | 386,704 |
| 13 | 7,502 | 1,160 | 8,662 | 379,202 |
| 14 | 7,525 | 1,138 | 8,662 | 371,677 |
| 15 | 7,547 | 1,115 | 8,662 | 364,130 |
| 16 | 7,570 | 1,092 | 8,662 | 356,560 |
| 17 | 7,593 | 1,070 | 8,662 | 348,967 |
| 18 | 7,615 | 1,047 | 8,662 | 341,352 |
| 19 | 7,638 | 1,024 | 8,662 | 333,713 |
| 20 | 7,661 | 1,001 | 8,662 | 326,052 |
| 21 | 7,684 | 978 | 8,662 | 318,368 |
| 22 | 7,707 | 955 | 8,662 | 310,661 |
| 23 | 7,730 | 932 | 8,662 | 302,930 |
| 24 | 7,754 | 909 | 8,662 | 295,177 |
| 25 | 7,777 | 886 | 8,662 | 287,400 |
| 26 | 7,800 | 862 | 8,662 | 279,600 |
| 27 | 7,824 | 839 | 8,662 | 271,776 |
| 28 | 7,847 | 815 | 8,662 | 263,929 |
| 29 | 7,871 | 792 | 8,662 | 256,059 |
| 30 | 7,894 | 768 | 8,662 | 248,164 |
| 31 | 7,918 | 744 | 8,662 | 240,246 |
| 32 | 7,942 | 721 | 8,662 | 232,305 |
| 33 | 7,965 | 697 | 8,662 | 224,339 |
| 34 | 7,989 | 673 | 8,662 | 216,350 |
| 35 | 8,013 | 649 | 8,662 | 208,337 |
| 36 | 8,037 | 625 | 8,662 | 200,299 |
| 37 | 8,061 | 601 | 8,662 | 192,238 |
| 38 | 8,086 | 577 | 8,662 | 184,152 |
| 39 | 8,110 | 552 | 8,662 | 176,042 |
| 40 | 8,134 | 528 | 8,662 | 167,908 |
| 41 | 8,159 | 504 | 8,662 | 159,749 |
| 42 | 8,183 | 479 | 8,662 | 151,566 |
| 43 | 8,208 | 455 | 8,662 | 143,359 |
| 44 | 8,232 | 430 | 8,662 | 135,126 |
| 45 | 8,257 | 405 | 8,662 | 126,869 |
| 46 | 8,282 | 381 | 8,662 | 118,588 |
| 47 | 8,307 | 356 | 8,662 | 110,281 |
| 48 | 8,332 | 331 | 8,662 | 101,949 |
| 49 | 8,357 | 306 | 8,662 | 93,593 |
| 50 | 8,382 | 281 | 8,662 | 85,211 |
| 51 | 8,407 | 256 | 8,662 | 76,805 |
| 52 | 8,432 | 230 | 8,662 | 68,373 |
| 53 | 8,457 | 205 | 8,662 | 59,915 |
| 54 | 8,483 | 180 | 8,662 | 51,433 |
| 55 | 8,508 | 154 | 8,662 | 42,925 |
| 56 | 8,534 | 129 | 8,662 | 34,391 |
| 57 | 8,559 | 103 | 8,662 | 25,832 |
| 58 | 8,585 | 77 | 8,662 | 17,247 |
| 59 | 8,611 | 52 | 8,662 | 8,636 |
| 60 | 8,636 | 26 | 8,662 | 0 |

#### Supplementary 27

Straight-Line GRT Amortization Table of tisagenlecleucel (Kymriah®) at an interest rate of 3% monthly over 10 years

| **Payment Period** | **Principal Amount Due** | **Interest Amount Due** | **Total Payment Amount Due** | **Principal Balance** |
| --- | --- | --- | --- | --- |
|  |  |  |  | 475,000 |
| 1 | 3,294 | 1,425 | 4,719 | 471,706 |
| 2 | 3,304 | 1,415 | 4,719 | 468,401 |
| 3 | 3,314 | 1,405 | 4,719 | 465,087 |
| 4 | 3,324 | 1,395 | 4,719 | 461,763 |
| 5 | 3,334 | 1,385 | 4,719 | 458,429 |
| 6 | 3,344 | 1,375 | 4,719 | 455,085 |
| 7 | 3,354 | 1,365 | 4,719 | 451,731 |
| 8 | 3,364 | 1,355 | 4,719 | 448,367 |
| 9 | 3,374 | 1,345 | 4,719 | 444,992 |
| 10 | 3,384 | 1,335 | 4,719 | 441,608 |
| 11 | 3,395 | 1,325 | 4,719 | 438,214 |
| 12 | 3,405 | 1,315 | 4,719 | 434,809 |
| 13 | 3,415 | 1,304 | 4,719 | 431,394 |
| 14 | 3,425 | 1,294 | 4,719 | 427,969 |
| 15 | 3,435 | 1,284 | 4,719 | 424,533 |
| 16 | 3,446 | 1,274 | 4,719 | 421,087 |
| 17 | 3,456 | 1,263 | 4,719 | 417,631 |
| 18 | 3,466 | 1,253 | 4,719 | 414,165 |
| 19 | 3,477 | 1,242 | 4,719 | 410,688 |
| 20 | 3,487 | 1,232 | 4,719 | 407,201 |
| 21 | 3,498 | 1,222 | 4,719 | 403,703 |
| 22 | 3,508 | 1,211 | 4,719 | 400,195 |
| 23 | 3,519 | 1,201 | 4,719 | 396,676 |
| 24 | 3,529 | 1,190 | 4,719 | 393,147 |
| 25 | 3,540 | 1,179 | 4,719 | 389,607 |
| 26 | 3,551 | 1,169 | 4,719 | 386,056 |
| 27 | 3,561 | 1,158 | 4,719 | 382,495 |
| 28 | 3,572 | 1,147 | 4,719 | 378,923 |
| 29 | 3,583 | 1,137 | 4,719 | 375,340 |
| 30 | 3,593 | 1,126 | 4,719 | 371,747 |
| 31 | 3,604 | 1,115 | 4,719 | 368,143 |
| 32 | 3,615 | 1,104 | 4,719 | 364,528 |
| 33 | 3,626 | 1,094 | 4,719 | 360,902 |
| 34 | 3,637 | 1,083 | 4,719 | 357,266 |
| 35 | 3,648 | 1,072 | 4,719 | 353,618 |
| 36 | 3,659 | 1,061 | 4,719 | 349,960 |
| 37 | 3,669 | 1,050 | 4,719 | 346,290 |
| 38 | 3,680 | 1,039 | 4,719 | 342,610 |
| 39 | 3,692 | 1,028 | 4,719 | 338,918 |
| 40 | 3,703 | 1,017 | 4,719 | 335,215 |
| 41 | 3,714 | 1,006 | 4,719 | 331,502 |
| 42 | 3,725 | 995 | 4,719 | 327,777 |
| 43 | 3,736 | 983 | 4,719 | 324,041 |
| 44 | 3,747 | 972 | 4,719 | 320,294 |
| 45 | 3,758 | 961 | 4,719 | 316,535 |
| 46 | 3,770 | 950 | 4,719 | 312,765 |
| 47 | 3,781 | 938 | 4,719 | 308,984 |
| 48 | 3,792 | 927 | 4,719 | 305,192 |
| 49 | 3,804 | 916 | 4,719 | 301,388 |
| 50 | 3,815 | 904 | 4,719 | 297,573 |
| 51 | 3,827 | 893 | 4,719 | 293,746 |
| 52 | 3,838 | 881 | 4,719 | 289,908 |
| 53 | 3,850 | 870 | 4,719 | 286,059 |
| 54 | 3,861 | 858 | 4,719 | 282,197 |
| 55 | 3,873 | 847 | 4,719 | 278,325 |
| 56 | 3,884 | 835 | 4,719 | 274,440 |
| 57 | 3,896 | 823 | 4,719 | 270,544 |
| 58 | 3,908 | 812 | 4,719 | 266,636 |
| 59 | 3,919 | 800 | 4,719 | 262,717 |
| 60 | 3,931 | 788 | 4,719 | 258,786 |
| 61 | 3,943 | 776 | 4,719 | 254,843 |
| 62 | 3,955 | 765 | 4,719 | 250,888 |
| 63 | 3,967 | 753 | 4,719 | 246,921 |
| 64 | 3,979 | 741 | 4,719 | 242,943 |
| 65 | 3,991 | 729 | 4,719 | 238,952 |
| 66 | 4,003 | 717 | 4,719 | 234,950 |
| 67 | 4,015 | 705 | 4,719 | 230,935 |
| 68 | 4,027 | 693 | 4,719 | 226,908 |
| 69 | 4,039 | 681 | 4,719 | 222,870 |
| 70 | 4,051 | 669 | 4,719 | 218,819 |
| 71 | 4,063 | 656 | 4,719 | 214,756 |
| 72 | 4,075 | 644 | 4,719 | 210,681 |
| 73 | 4,087 | 632 | 4,719 | 206,594 |
| 74 | 4,100 | 620 | 4,719 | 202,494 |
| 75 | 4,112 | 607 | 4,719 | 198,382 |
| 76 | 4,124 | 595 | 4,719 | 194,258 |
| 77 | 4,137 | 583 | 4,719 | 190,122 |
| 78 | 4,149 | 570 | 4,719 | 185,973 |
| 79 | 4,161 | 558 | 4,719 | 181,811 |
| 80 | 4,174 | 545 | 4,719 | 177,637 |
| 81 | 4,186 | 533 | 4,719 | 173,451 |
| 82 | 4,199 | 520 | 4,719 | 169,252 |
| 83 | 4,212 | 508 | 4,719 | 165,040 |
| 84 | 4,224 | 495 | 4,719 | 160,816 |
| 85 | 4,237 | 482 | 4,719 | 156,579 |
| 86 | 4,250 | 470 | 4,719 | 152,329 |
| 87 | 4,262 | 457 | 4,719 | 148,067 |
| 88 | 4,275 | 444 | 4,719 | 143,792 |
| 89 | 4,288 | 431 | 4,719 | 139,504 |
| 90 | 4,301 | 419 | 4,719 | 135,203 |
| 91 | 4,314 | 406 | 4,719 | 130,889 |
| 92 | 4,327 | 393 | 4,719 | 126,562 |
| 93 | 4,340 | 380 | 4,719 | 122,223 |
| 94 | 4,353 | 367 | 4,719 | 117,870 |
| 95 | 4,366 | 354 | 4,719 | 113,504 |
| 96 | 4,379 | 341 | 4,719 | 109,126 |
| 97 | 4,392 | 327 | 4,719 | 104,734 |
| 98 | 4,405 | 314 | 4,719 | 100,328 |
| 99 | 4,418 | 301 | 4,719 | 95,910 |
| 100 | 4,432 | 288 | 4,719 | 91,478 |
| 101 | 4,445 | 274 | 4,719 | 87,033 |
| 102 | 4,458 | 261 | 4,719 | 82,575 |
| 103 | 4,472 | 248 | 4,719 | 78,104 |
| 104 | 4,485 | 234 | 4,719 | 73,618 |
| 105 | 4,499 | 221 | 4,719 | 69,120 |
| 106 | 4,512 | 207 | 4,719 | 64,608 |
| 107 | 4,526 | 194 | 4,719 | 60,082 |
| 108 | 4,539 | 180 | 4,719 | 55,543 |
| 109 | 4,553 | 167 | 4,719 | 50,991 |
| 110 | 4,566 | 153 | 4,719 | 46,424 |
| 111 | 4,580 | 139 | 4,719 | 41,844 |
| 112 | 4,594 | 126 | 4,719 | 37,250 |
| 113 | 4,608 | 112 | 4,719 | 32,643 |
| 114 | 4,621 | 98 | 4,719 | 28,021 |
| 115 | 4,635 | 84 | 4,719 | 23,386 |
| 116 | 4,649 | 70 | 4,719 | 18,737 |
| 117 | 4,663 | 56 | 4,719 | 14,074 |
| 118 | 4,677 | 42 | 4,719 | 9,396 |
| 119 | 4,691 | 28 | 4,719 | 4,705 |
| 120 | 4,705 | 14 | 4,719 | 0 |
